# Altered Network Efficiency in Isolated REM Sleep Behaviour Disorder: A Multicentric Study

**DOI:** 10.1101/2025.07.08.25331130

**Authors:** Christina Tremblay, Alexandre Pastor-Bernier, François Rheault, Véronique Daneault, Violette Ayral, Marie Filiatrault, Liane Desaulniers, Andrew Vo, Jean-François Gagnon, Ronald B. Postuma, Petr Dusek, Stanislav Marecek, Zsoka Varga, Johannes C. Klein, Michele T. Hu, Stéphane Lehéricy, Isabelle Arnulf, Pauline Dodet, Marie Vidailhet, Jean-Christophe Corvol, ICEBERG Study Group, Shady Rahayel

**Affiliations:** Centre for Advanced Research in Sleep Medicine, Hôpital du Sacré-Cœur de Montréal, Montreal, QC, Canada, H4J 1C5; Sherbrooke Connectivity Imaging Lab, Université de Sherbrooke, Sherbrooke, QC, Canada, J1K 0A5; Department of Neuroscience, University of Montreal, Montreal, QC, Canada H3C 3J7; The Neuro (Montreal Neurological Institute), McGill University, Montreal, QC, Canada, H3A 2B4; Department of Psychology, Université du Québec à Montréal, Montreal, QC, Canada, H2X 3P2; Research Centre, Institut universitaire de gériatrie de Montréal, Montreal, QC, Canada, H3W 1W5; Department of Neurology, Montreal General Hospital, Montreal, QC, Canada, H3G 1A4; Department of Neurology and Centre of Clinical Neurosciences, First Faculty of Medicine, Charles University and General University Hospital, Prague, Czechia, 121 08; Oxford Parkinson’s Disease Centre and Division of Neurology, Nuffield Department of Clinical Neurosciences, University of Oxford, Oxford, UK, 0X3 7JX; Institut du Cerveau (Paris Brain Institute, ICM), Sorbonne Université, Inserm, CNRS, Assistance Publique Hôpitaux de Paris, Paris, France, 75013; Department of Medicine, University of Montreal, Montreal, QC, Canada H3C 3J7

**Keywords:** structural connectivity, graph theory, diffusion MRI, synucleinopathies, sleep, parasomnias

## Abstract

Isolated REM sleep behavior disorder (iRBD), a prodromal synucleinopathy, generally precedes Parkinson’s disease (PD) or dementia with Lewy bodies (DLB). While disruptions in structural brain connectivity have been reported in these diseases, their presence in prodromal phases such as iRBD remains unclear. In this cross-sectional study, we analysed diffusion MRI in a large multicentric dataset (198 iRBD, 174 controls), mapping white matter pathways between 462 regions. Comparing groups, we found disrupted structural connectivity in iRBD. This included reduced density in multiple cortical areas alongside focal increases suggesting compensation in parietal, orbitofrontal and visual cortices. Global and local efficiency were altered in iRBD, notably in motor-related regions (putamen, thalamus, sensorimotor and parietal cortices), and was associated with emerging motor features. Importantly, increased local efficiency in the supramarginal gyrus predicted phenoconversion to DLB, but not PD. These findings highlight early structural connectivity disruptions in iRBD, offering a potential marker for progression to DLB.

## Introduction

Isolated REM sleep behavior disorder (iRBD) is a parasomnia characterized by abnormal movements during REM sleep. Over 80% of individuals with iRBD develop a synucleinopathy within 10 to 15 years, most commonly Parkinson’s disease (PD) or dementia with Lewy bodies (DLB). This makes iRBD the strongest known prodromal predictor for these conditions, offering a critical window to investigate the early structural brain changes preceding synucleinopathies and potentially identify therapeutic targets.

White matter damage and altered structural connectivity, detectable via diffusion-weighted imaging (DWI), are recognized features in PD and DLB.^4,5^ Meta-analyses confirm widespread white matter alterations primarily affecting subcortical, limbic, and cortical regions, even in early PD stages.^6–8^ These disruptions impact network organization, leading to poorer integration and segregation, characterized by decreased clustering coefficient (regions less closely interconnected in local clusters) and lower global efficiency (less efficient information transfer).^4^ Similarly, structural connectivity alterations are reported in DLB, though findings vary regarding their extent.^5^ Some studies reported widespread white matter disruptions^9^ while others found disruptions confined to parietal and occipital regions.^10^ Several studies identified increased network efficiency and connectivity in DLB,^11–13^ interpreted as a compensatory mechanism that may become burdensome as pathology progresses.^14^ How and when connectivity changes occur remain to be understood in both PD and DLB.

Alterations in white matter and structural connectivity in iRBD have gained attention as potential indicators for prodromal PD or DLB. Initial work with 20 prodromal PD (with iRBD and/or hyposmia) found increase connectivity in the supplementary motor area, parahippocampal gyrus, putamen and cerebellum, potentially reflecting compensatory mechanisms.^15^ Others found decreased global and local network efficiency, especially in frontal cortex, lower degree (lower number of connections) and increased eigenvector centrality (stronger influence of regions within the network) in the caudate and frontal cortex in 10 iRBD.^16^ These findings suggest network alterations in iRBD, but larger cohorts are needed to more reliably identify the early structural connectivity changes.

Therefore, this study leverages a large, international, multicentric DWI dataset from polysomnography-confirmed iRBD participants (n=198) and healthy controls (n=174). We aimed to characterize structural connectivity alterations using network-based statistics (NBS) for connection density and graph theory to assess global and local network efficiency. We further examined relationships between local efficiency and other network metrics (clustering coefficient, connection strength, degree and eigenvector centrality) to define network reorganization. Finally, we investigated whether efficiency changes predict phenoconversion towards DLB versus PD in the 177 iRBD patients followed longitudinally. We hypothesized that iRBD patients would show localized reduced connection density along with lower global and local network efficiency compared to controls.

## Results

### Participants

The iRBD group had worse global cognition and parkinsonian motor features compared to the control group as indicated by their MoCA and MDS-UPDRS-III scores, respectively (Table 1).

**Table 1.** Demographics, clinical variables and graph theory measures of participants with iRBD compared to controls.

| Variables | iRBD (n=198) | Controls (n=174) | T-test |
| --- | --- | --- | --- |
|  | mean (SD) | mean (SD) | P-value |
| Sex (men/women)<sup>a</sup> | 174/24 | 99/75 | < **0.001** |
| Age (years)<sup>b</sup> | 66.5 (7.0) | 66.2 (6.9) | 0.64 |
| MoCA<sup>b</sup> | 25.6 (3.0) | 27.0 (2.4) | < **0.001** |
| MDS-UPDRS-III<sup>b</sup> | 6.4 (5.6) | 3.9 (7.1) | **0.001** |
| Global efficiency (W-score)<sup>c</sup> | -0.31 (1.04) | NA | < **0.00001** |
| Local efficiency (W-score)<sup>c</sup><br>low-efficiency regions | -0.28(0.08) | NA | < **0.00001** |
| Local efficiency (W-score)<sup>c</sup><br>high-efficiency regions | 0.27 (0.04) | NA | < **0.00001** |
| Clustering coefficient (W-score)<sup>c</sup><br>low-efficiency regions | -0.30 (0.09) | NA | < **0.00001** |
| Clustering coefficient (W-score)<sup>c</sup><br>high-efficiency regions | 0.26 (0.06) | NA | < **0.00001** |
| Connection strength (W-score)<sup>c</sup><br>low-efficiency regions | 0.06 (0.13) | NA | 0.06 |
| Connection strength (W-score)<sup>c</sup><br>high-efficiency regions | -0.30 (0.11) | NA | **0.00004** |
| Degree (W-score)<sup>c</sup><br>low-efficiency regions | 0.06 (0.13) | NA | 0.04 |
| Degree (W-score)<sup>c</sup><br>high-efficiency regions | -0.31 (0.10) | NA | **0.00002** |
| Eigenvector centrality (W-score)<sup>c</sup><br>low-efficiency regions | 0.01 (0.11) | NA | 0.71 |
| Eigenvector centrality (W-score)<sup>c</sup><br>high-efficiency regions | -0.06 (0.11) | NA | 0.19 |

Regions with significant alterations in local efficiency were separate in low- (negative W-score) vs. High-efficiency regions (positive W-score)
<sup>a</sup>χ<sup>2</sup> test
<sup>b</sup> Two-sample t-test (Student's t-test)
<sup>c</sup> One-sample t-test
MoCA: Montreal Cognitive Assessment; MDS: Movement Disorders Society;
UPDRS-III: Unified Parkinson's Disease Rating Scale part.3 (motor scale); NA: Not applicable

### Structural connectivity is altered in iRBD

Structural connectivity maps were analyzed to assess significant regional changes in connectivity density in iRBD patients compared to controls (Fig.1A and 1B). Network-based statistics revealed reduced structural connectivity in iRBD relative to controls in 14 connections across 27 different regions (*p-value_FDR_* < 0.05) (Supplementary Table 1). Reduced connectivity in the left hemisphere were observed in frontal (precentral, paracentral, parsorbitalis, rostral middle frontal, superior frontal, lateral orbitofrontal and, rostral and caudal anterior cingulate cortices) and postcentral regions. In the right hemisphere, reductions were predominantly found in the frontal (medial orbitofrontal, superior frontal and precentral gyrus), temporal and posterior regions, including the fusiform, lingual, inferior parietal, posterior cingulate and lateral occipital cortices. Increased connectivity was also found in three connections between the right inferior and superior parietal cortex, and in one connection between the left medial orbitofrontal and visual cortex (*p-value_FDR_* < 0.05). The number of regions with statistically significant changes did not differ between the two hemispheres: right (21/231 = 9% of regions) compared to the left (12/231 = 5% of regions) hemisphere (c^2^ = 2.64; *p-value* = 0.10).

**Figure 1.**
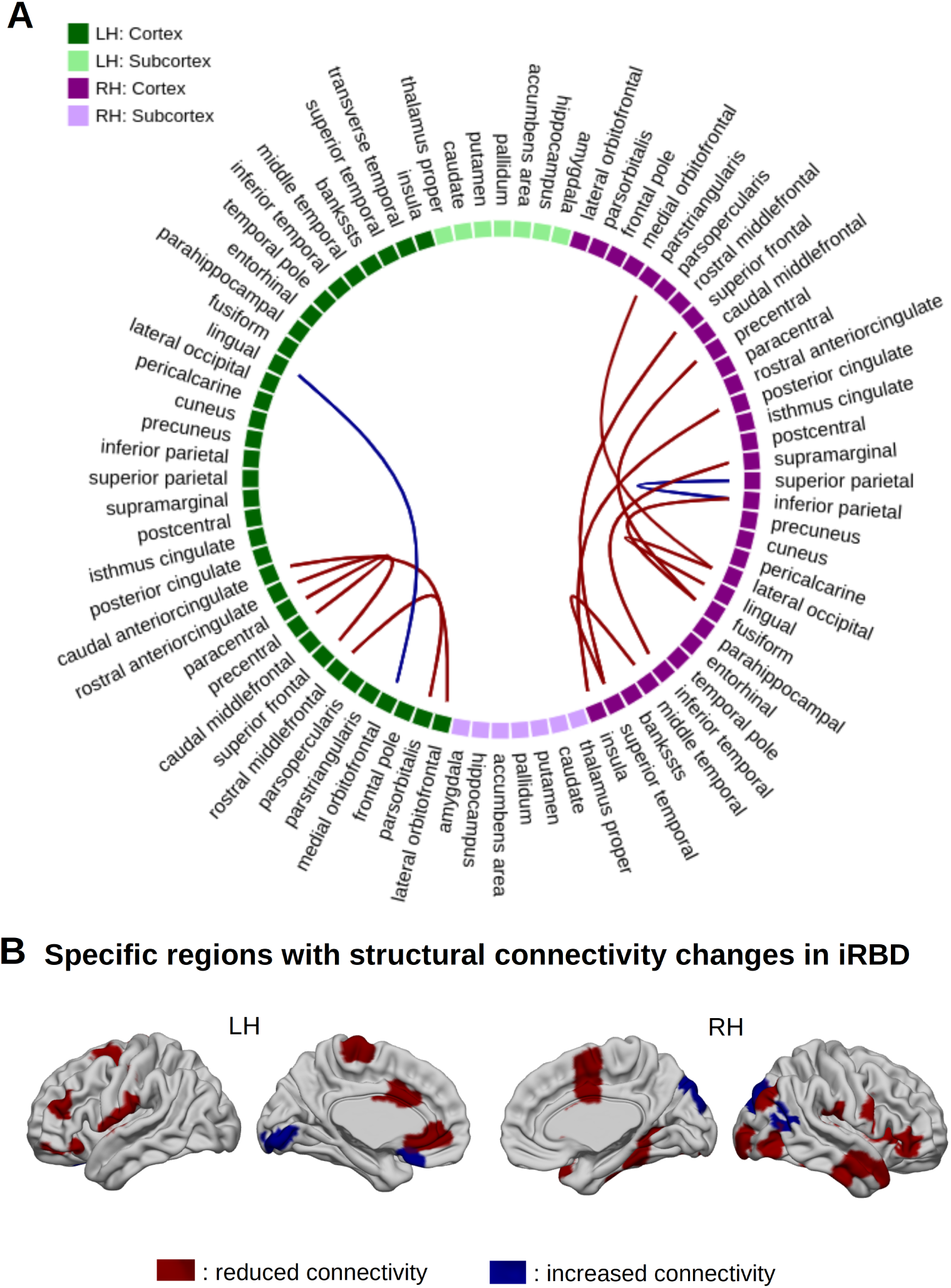
Structural connectivity changes in patients with isolated REM sleep behavior disorder (iRBD) (**A**) Circular plot showing the structural connections with significantly lower (red) and higher (blue) connection density in iRBD compared to controls (*p-value_FDR_*<.05 using network-based statistics with age, sex, and center as covariates). Subregions of the Cammoun atlas have been combined in 78 regions for visualization. (**B**) Brain map representations of the 27 subregions showing significantly lower connectivity (red areas) and the 6 subregions with stronger connectivity (blue areas) in iRBD compared to controls. FDR = False Discovery Rate; iRBD = isoled REM sleep behavior disorder

### Structural network efficiency is altered in iRBD

To assess whether connectivity changes impacted network efficiency at global and local scales, we quantified the global efficiency of each subject’s connectome as well as the local efficiency of each region. This analysis provided insights into how connectivity changes influence network efficiency. To isolate disease-specific effects from those related to aging and sex, global and local efficiency W-scores were used. A significant decrease in global efficiency was observed in iRBD (W-score = -0.31, *p-value* < 0.001; Supplementary Fig.1) compared to controls. Furthermore, local efficiency was reduced in 18 cortical and three subcortical (left putamen and bilateral thalamus) regions and increased in 11 cortical regions after FDR corrections (Fig.2A, Supplementary Table 2). Regions showing the largest reduction in local efficiency were the postcentral cortex (W-score = -0.52), putamen (-0.41), inferior parietal cortex (-0.41), thalamus (-0.37), and precentral cortex (-0.33), all involved in motor functioning. Interestingly, no regions showed changes in local efficiency and connection density simultaneously. Taken together, these findings suggest that both specific connections and efficiency changes occur in iRBD and might contribute to reduced brain global efficiency.

**Figure 2.**
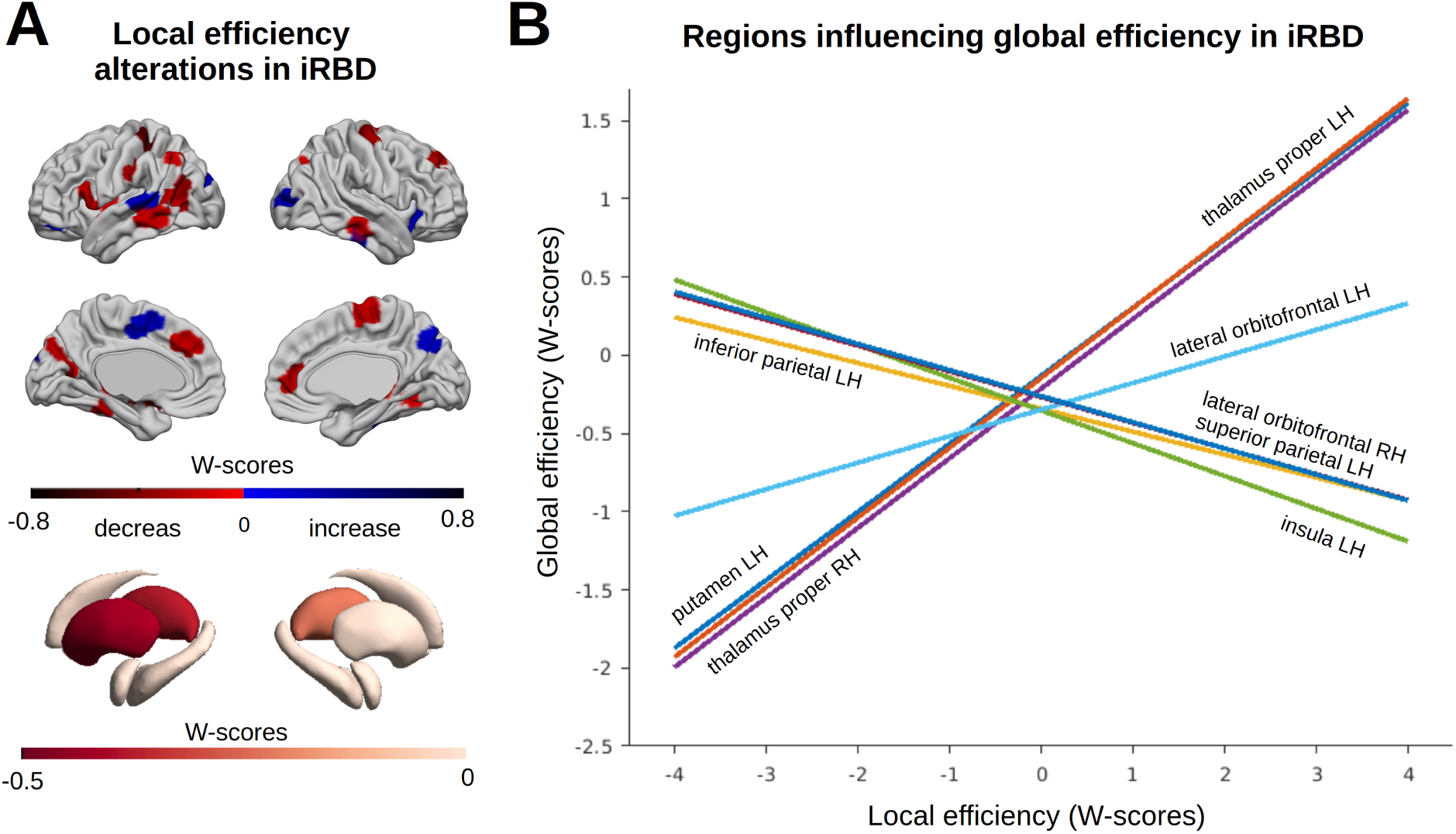
Local efficiency alterations in participants with isolated REM sleep behavior disorder (iRBD) (**A**) Cortical and subcortical regions showing significant decreases (red) and increases (blue) in local efficiency in the iRBD group compared to controls (*p-value_FDR_*<.05). (**B**) Decrease and increase in local efficiency in specific cortical and subcortical regions are associated with the global efficiency reduction observed in iRBD. RH: Right hemisphere; LH: Left hemisphere

### Subcortical and cortical regions differentially contribute to impaired global efficiency

To identify the regions contributing to the reduction in global efficiency observed in iRBD, a stepwise regression analysis was performed. The local efficiency W-scores of the 30 regions with significant changes in iRBD were used as predictors. Eight regions were identified as significant contributors to global efficiency changes in iRBD. Four regions, namely the left putamen, left and right thalamus, and left lateral orbitofrontal gyrus, were positive contributors. This indicates that lower local efficiency in these regions was associated with lower global efficiency. In contrast, four other regions, the left inferior and superior parietal cortex, left insula and right orbitofrontal cortex, were negative contributors, indicating that higher local efficiency in these regions was associated with lower global efficiency (Fig.2B and Supplementary Table 3). These findings indicate that decreased local efficiency, notably in subcortical regions, directly contributes to the reduction in global network efficiency. Conversely, increased local efficiency in specific cortical regions contributes to the decrease in global efficiency, suggesting that these increases occur at the expense of global efficiency in iRBD.

### Regions with altered efficiency have specific topological network characteristics

To further understand how changes in network organization impact local efficiency in iRBD, we analyzed the relationships between local efficiency and other graph measures, namely clustering coefficient, connection strength, degree and eigenvector centrality. W-scores were used to quantify the deviation in iRBD patients from controls. One-sample t-tests with FDR correction were applied to the W-scores for clustering coefficient, connection strength, degree and eigenvector centrality to identify significant differences between groups. A significant reduction was observed in 48 regions in clustering coefficient, while nine regions showed increased clustering (Supplementary Table 4, Fig.3A). Connection strength was significantly decreased in 102 regions while it was increased in eight regions, predominantly in the frontal lobe as well as in the fusiform, lingual, postcentral and inferior parietal cortex (Supplementary Table 5, Fig.3A). In addition, lower degree (number of connections) was found in iRBD in 105 regions while seven regions had higher degree, each also showing increased connection strength (Supplementary Table 6, Fig.3A). There was no significant difference in eigenvector centrality compared to controls.

**Figure 3.**
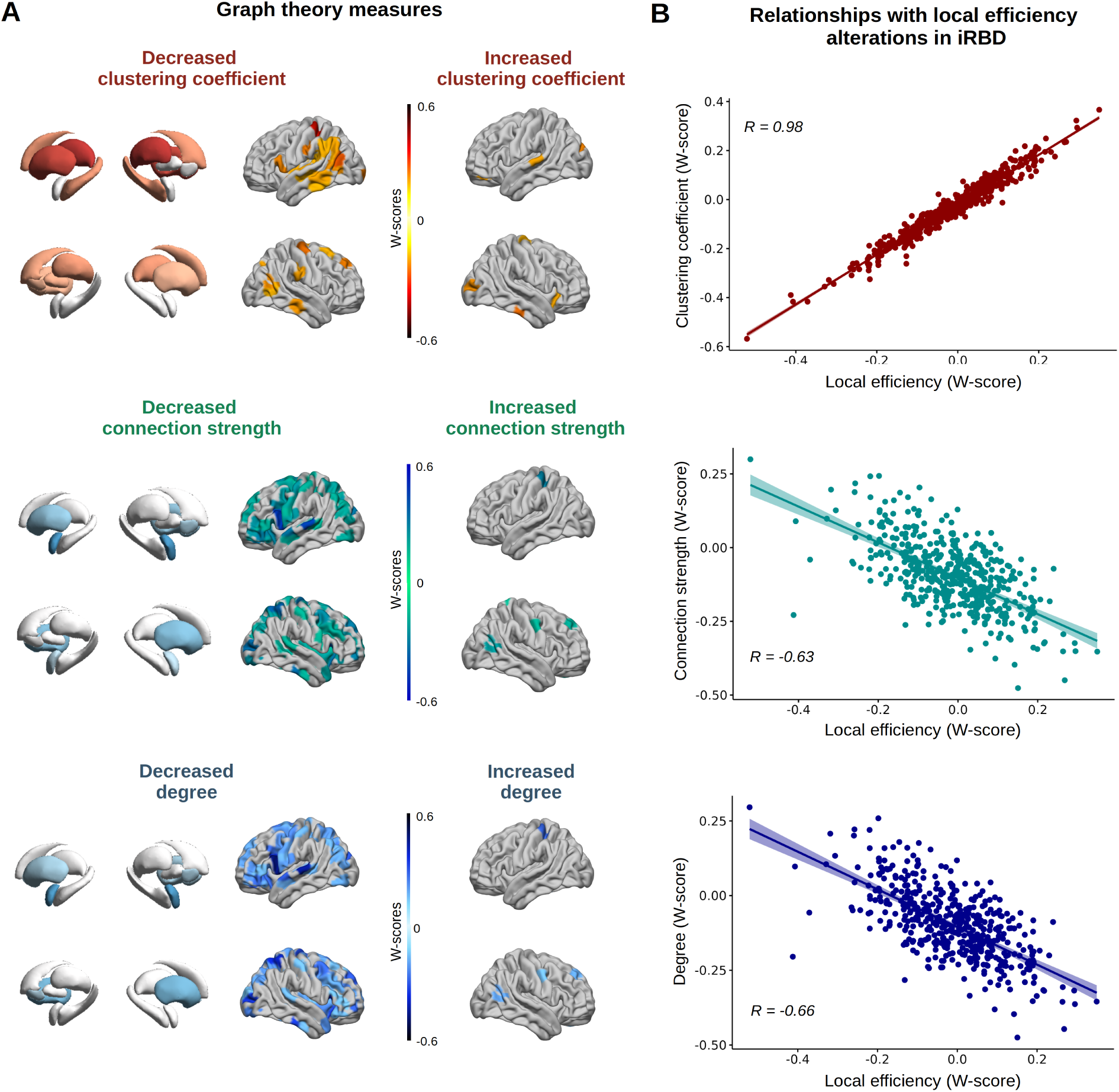
Relationships between local efficiency alterations in the isolated REM sleep behavior disorder (iRBD) group and graph theory metrics. (**A**) Subcortical and cortical brain maps illustrating significant regional differences in iRBD compared to controls (W-scores) for clustering coefficient, connection strength and degree (number of connections). (**B**) Significant positive spatial correlations were found between local efficiency alterations in iRBD and the clustering coefficients while significant negative correlations were observed between local efficiency and both connection strength and degree in iRBD. All correlations (Pearson’s *r*) were compared against null coefficient distributions using a model that preserves spatial autocorrelation between regions, with FDR correction (significance: *p-value_spin-FDR_*<.05).

Regions with the largest reduction in clustering were the postcentral gyrus (W-score=-0.57), inferior parietal cortex (-0.42), thalamus (-0.42), putamen (-0.39), supramarginal (-0.34), and precentral cortex (-0.33). Among the 48 regions with decreased clustering, 21 (44%) also showed reduced efficiency. Of the nine regions with increased clustering, six regions (67%) also exhibited increased efficiency. Interestingly, all regions with reduced local efficiency in iRBD had decreased clustering coefficient. This suggests that reduced clustering might be a precursor or primary condition for the alterations in local efficiency in iRBD. Spatial Pearson correlations revealed a positive correlation between local efficiency in each of the 462 regions and clustering coefficient (*r* = 0.98, *p-value_spin-FDR_* = 0.001) and negative correlations with connection strength (*r* = -0.63, *p-value_spin-FDR_*= 0.001) and degree (*r* = -0.66, *p-value_spin-FDR_* = 0.001) (Fig.3B). No significant correlation was observed with eigenvector centrality (*r* = -0.02, *p-value_spin-FDR_* = 0.38).

Next, we investigated whether regions with higher (significant positive) and lower (significant negative) local efficiency W-scores in iRBD differed based on the graph theory metrics. Consistent with the correlation results, independent samples t-tests indicated significant differences between regions with higher and lower local efficiency in terms of clustering coefficient [t(28) = -17.51, *p-value_FDR_* < 0.00001], connection strength [t(28) = 7.07, *p-value_FDR_* < 0.00001], and degree [t(28) = 7.40, *p-value_FDR_* < 0.00001]. However, no difference was observed in eigenvector centrality [t(28) = 1.42, *p-value_FDR_*= 0.17]. It is noteworthy that there were no significant correlations in regions with lower local efficiency (N=21) between local efficiency and degree (*r* = -0.18, *p-value_spin_* = 0.20) or connection strength (r = -0.18, *p-value_spin_* = 0.21), but a positive correlation was observed with the clustering coefficient (r = 0.90, *p-value_spin_*= 0.001). These results indicate that higher-efficiency regions in iRBD are more clustered with neighboring regions despite showing fewer and weaker connections, potentially reflecting greater tissue damage, while low-efficiency regions are less clustered with neighboring regions.

### Local efficiency associates with parkinsonian motor features

We also investigated whether local efficiency relates to cognitive and motor features in iRBD patients using PLS regression between local efficiency (absolute W-scores) and the MoCA and MDS-UPDRS-III scores. A total of 186 iRBD patients with a MoCA score and 175 iRBD patients with an MDS-UPDRS-III score were included in these analyses. Of the 10 latent variables tested, we identified one significant latent variable where local efficiency associated with MDS-UPDRS-III scores (*p-value* = 0.03). This latent variable accounted for 19.5% of the covariance between local efficiency and parkinsonian motor features (Supplementary Fig.2). Bootstrapping identified eight regions robustly associated with this latent variable, with ratios ranging from 4.0 to 5.5 (p<0.0001), namely the right superior parietal cortex (two subregions), insula, inferior parietal and medial orbitofrontal cortex, as well as the left lateral occipital, lateral orbitofrontal, and inferior parietal cortex. In three of these regions, higher changes in local efficiency were correlated with increased MDS-UPDRS-III scores (right insula: *r* = -0.24, *p-value* = 0.001, right medial orbitofrontal cortex: *r* = -0.20, *p-value* = 0.007, and left lateral occipital cortex: *r* = -0.16, *p-value* = 0.04) (Supplementary Fig.3). In contrast, no significant latent variables were identified in relation with the MoCA. These findings suggest that emerging parkinsonian motor features in iRBD primarily covary with efficiency changes in insular, orbitofrontal, and occipital regions.

### Supramarginal gyrus local efficiency predicts DLB but not PD phenoconversion

Linear regression analyses controlling for age, sex and follow-up time were conducted to assess whether there are local efficiency (W-scores) differences between individuals who converted to PD or DLB, and those who remained disease-free (non-converters). Among the 462 brain regions assessed, only the right supramarginal gyrus (region 105) showed a significant increase in local efficiency in DLB converters compared to non-converters (mean W-score: 1.35 ± 1.45 vs. –0.10 ± 1.09; estimate = 1.88, t = 4.63, *p-value_FDR_*= 0.005). No regions were significantly different in PD converters.

Time-to-event analyses found that the local efficiency of the right supramarginal gyrus was a strong and significant predictor of DLB conversion over time (β = 0.90, z = 3.66, *p-value* = 0.0002), with a hazard ratio (HR) of 2.45 (95% CI: 1.52–3.97). This indicates that each 1-point increase in local efficiency of the right supramarginal gyrus was associated with a 2.5-fold increase in the hazard of converting to DLB, independent of age and sex. Neither age (HR = 1.08, *p-value* = 0.18) nor sex (HR = 0.26, *p-value* = 0.14) reached statistical significance in the presence of local efficiency. In contrast, the W-score local efficiency of the right supramarginal gyrus was not a significant predictor of PD conversion (β = 0.24, z = 1.56, *p-value* = 0.12), with a hazard ratio of 1.27 (95% CI: 0.94–1.72). Neither age (HR = 1.02, *p-value* = 0.63) nor sex (HR = 7.56×10⁷, *p-value* = 0.998) were significant in the PD model. Kaplan-Meier survival curves were used to compare survival rates between iRBD patients showing positive versus negative W-score local efficiency in the right supramarginal gyrus. They demonstrated that almost all iRBD patients with a negative local efficiency did not convert to DLB during follow-up, in contrast to those with positive local efficiency (Fig.4). These results support the clinical relevance of local efficiency for the prognosis of DLB in iRBD.

**Figure 4.**
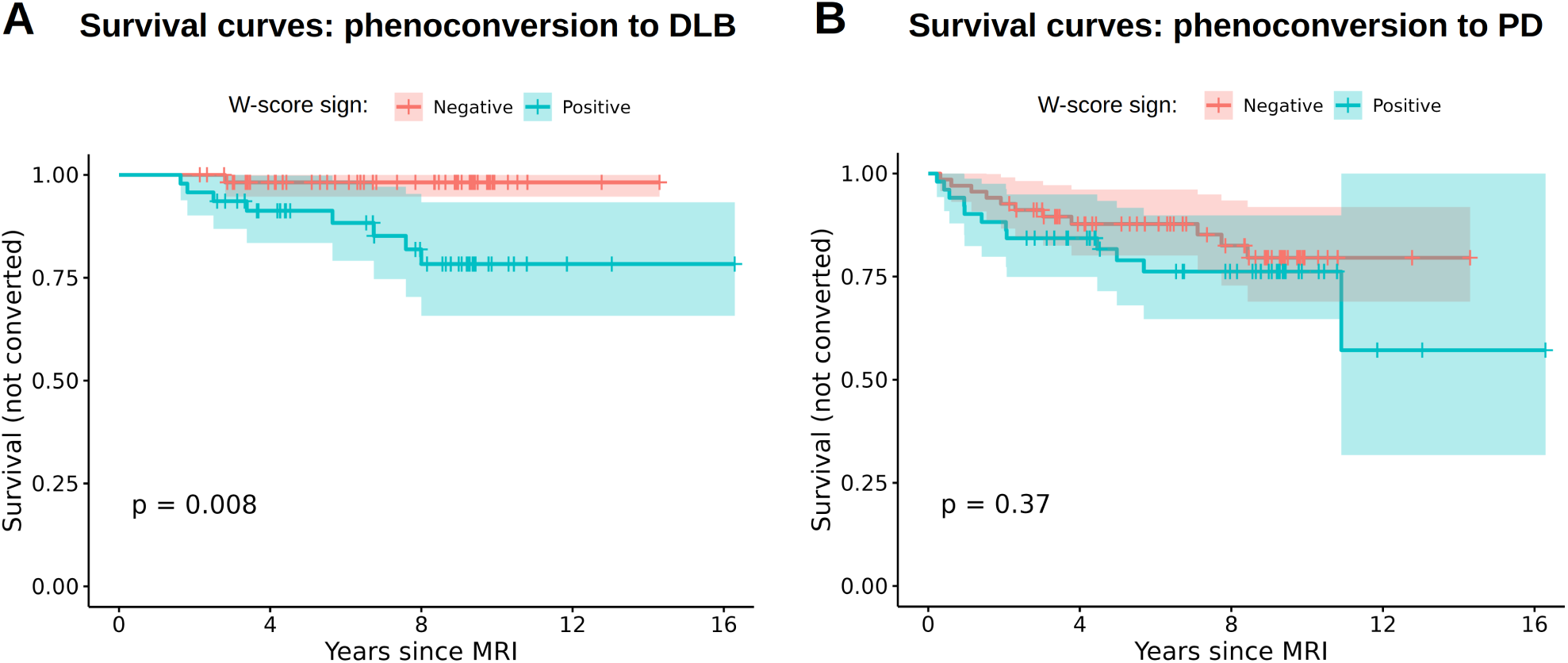
Kaplan–Meier survival curves showing the association between local efficiency in the right supramarginal gyrus (W-score) and phenoconversion in isolated REM sleep behavior disorder (iRBD) **(A)** Time to phenoconversion in DLB. Individuals with iRBD with a positive W-score (pink line) in the right supramarginal gyrus showed a significantly lower survival probability (i.e., higher risk of conversion), compared to those with a negative W-score (blue line); **(B)** Time to phenoconversion in PD. No significant difference in survival was observed between individuals with iRBD with positive (pink) and negative (blue) W-scores. P-value (p) indicates statistical significance from the log-rank test.

## Discussion

Leveraging one of the largest iRBD diffusion MRI datasets, we identified different structural connectivity alterations and their impact on network efficiency. Our findings reveal distinct structural disconnection patterns in iRBD, with the most pronounced disruptions in frontal and posterior brain regions. Both global and local network efficiency were significantly reduced, especially in subcortical and sensorimotor areas, suggesting widespread alterations consistent with early synucleinopathy. Regions with higher local efficiency were more clustered but less strongly connected. Importantly, efficiency changes correlated with motor features and predicted DLB conversion, demonstrating ongoing network reorganization related to disease progression.

Network-Based Statistics identified reduced connectivity in 14 bilateral connections involving all major lobes, particularly frontal, temporal, and posterior regions, including the insula. These findings align with previous studies reporting widespread structural alterations in iRBD.^17–19^ Widespread atrophy was reported across the frontal, inferior temporal, parietal and occipital cortices.^17^ Disrupted connectivity in the insula, and middle frontal cortex has also been observed in iRBD and PD.^20^ In our study, reduced connectivity density was similarly found in these regions, along with additional frontal and posterior areas. Differences with previous studies may reflect the use of a higher resolution parcellation, larger sample and the use of Convex Optimization Modeling for Microstructure Informed Tractography (COMMIT2) for tractography to minimize false positive connections.^21^

The iRBD group showed decreased global efficiency and reduced local efficiency in 18 cortical and three subcortical regions: the left putamen and bilateral thalamus, critical for motor, cognitive and sensory functions (Fig.5A).^22–24^ Reduced efficiency in the putamen aligns with the dopaminergic input loss, associated with motor symptoms in DLB and PD,^25^ as well as iRBD studies showing decreased putamen volume.^26,27^ Similarly, atrophy in the thalamus has been reported in iRBD patients with mild cognitive impairment.^28^ Conversely, increased local efficiency was found in nine cortical regions across the frontal, temporal, parietal, and occipital lobes (Fig.5B). These regions had higher clustering coefficients but fewer and weaker connections. In this study, clustering coefficient was reduced in 48 regions and increased in nine regions. All 21 regions with reduced local efficiency had decreased clustering, suggesting that changes in clustering may precede or drive local efficiency alterations in iRBD. Regions with increased efficiency may be undergoing reorganization, potentially pruning weak connections to preserve localized function.^29,30^ This process, while possibly beneficial in the short term, may impair global communication efficiency.

**Figure 5.**
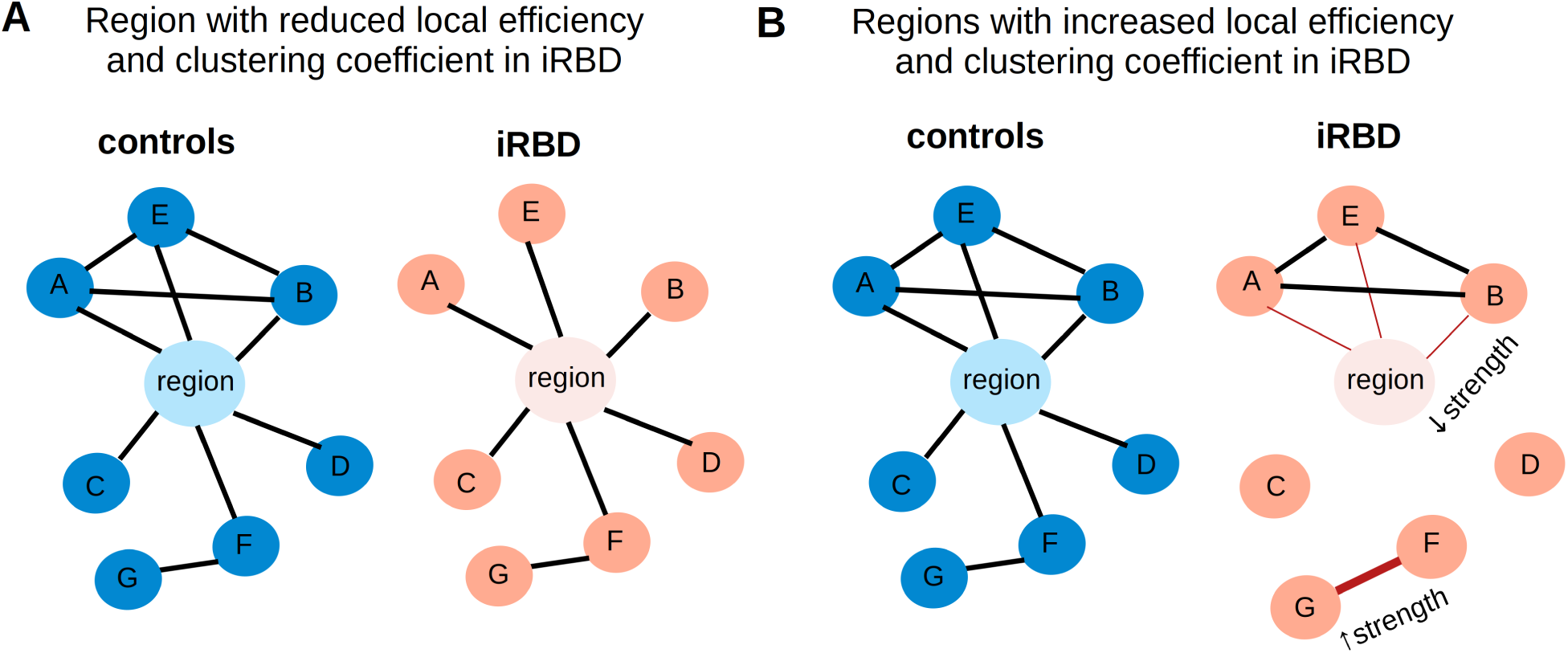
Schematic representation of potential network alterations in isolated REM sleep behavior disorder (iRBD) (**A**) Example of network disruption in iRBD compared to controls, where neighboring regions (A, B, E) lose connections (black lines) between them, resulting in reduced local efficiency and clustering coefficient in the central region (**B**) Example of network reorganization leading to increase local efficiency and clustering coefficient along with lower degree (number of direct connections) and average connection strength (thinner red lines) in a region. Possibly due to loss connections and inputs, leading to reduce competition, increase strength (or density) in specific connection (thicker red line) can happen among more isolated neighbor regions (G and F).

Importantly, changes in network topology were clinically relevant. Local efficiency in the insula, orbitofrontal, and occipital cortices was associated with parkinsonian motor features (MDS-UPDRS-III). While these regions are not primary motor areas, they are affected early in synucleinopathies^31,32^ and are associated with symptoms such as olfactory loss,^32,33^ cognitive decline,^34^ color discrimination and autonomic dysfunctions.^28,31^ Our findings support the view that motor symptom scores in iRBD may reflect broader disease severity rather than isolated motor impairment.

We also identified increased connectivity between the right superior and inferior parietal cortices, and between the left medial orbitofrontal and visual cortex. These patterns may represent compensatory hyperconnectivity in response to early degeneration, consistent with prior reports in prodromal PD and DLB.^13,15^ While such increases may initially help maintain function, chronic over-recruitment of affected networks could become inefficient over time and contribute to functional decline.^35^ Notably, increased local efficiency in the right supramarginal gyrus (area 105) predicted conversion to DLB but not PD. While this region did not differ between iRBD and controls in group-level efficiency, other sub-regions of the supramarginal gyrus showed reduced connectivity strength (area 104), clustering (area 109) and number of connections (area 104). This suggests that the increase in local efficiency may reflect disrupted intra-regional connectivity. Given the known involvement of parietal regions in DLB, including white matter damage^36,37^ and posterior hypoperfusion^38^, local efficiency in the right supramarginal gyrus may serve as a potential marker of risk for DLB.

This study has several strengths. It includes a large sample of polysomnography-confirmed iRBD patients and controls with harmonized acquisition, improving generalizability and enabling detection of more subtle differences compared to previous study. This study also used COMMIT2 for tractogram construction, which improves accuracy by filtering out false-positive streamlines that commonly inflate connectivity estimates. We also used Tractoflow-ABS, which incorporates atlas-based segmentation, FreeSurfer priors, and accounts for atrophy to further improve tract accuracy. Finally, we choose a high-resolution atlas (462 regions), to enable detection of a more detailed set of brain connections compared to standard atlases like Desikan-Killiany and Desikan-Killiany-Tourville atlases. However, some limitations remain. First, our analyses focused exclusively on structural connectivity. Incorporating resting-state functional MRI measures would clarify whether these structural disruptions lead to functional impairments. Second, clinical assessments were limited to MDS-UPDRS-III and MoCA, and could not fully capture the phenotypic heterogeneity of iRBD. Third, although sex was adjusted for, the iRBD sample had a high men predominance, limiting sex-specific interpretations. Future studies should incorporate more women and further explore how sex interacts with network alterations.

In conclusion, our results provide robust evidence for early structural network reorganization in iRBD, marked by reduced global efficiency and region-specific alterations in local efficiency. Notably, subcortical and sensorimotor regions showed marked reductions in local efficiency, contributing to lower global efficiency, consistent with synucleinopathy-related neurodegeneration. Clinically, local efficiency alterations were associated with motor features and predicted DLB conversion. These findings identify candidate imaging markers for prodromal synucleinopathies and underscore the value of connectivity approaches to track disease progression and stratify risk in iRBD.

## Methods

### Participants

Clinical and neuroimaging data, including T1-weighted MRI and DWI, from individuals with iRBD and healthy controls were acquired by different international centers including Canada (Montreal: two scanners), Czech Republic (Prague: one scanner), United Kingdom (Oxford: one scanner) and France (Paris: two scanners) (see Supplementary Table 7). Baseline data from iRBD and controls in the Parkinson’s Progression Markers Initiative (PPMI, http://www.ppmi-info.org), a longitudinal multicenter study with standardized acquisition protocols, were added to the dataset.^39^ Finally, controls data in the Quebec Parkinson Network initiative (QPN; https://rpq-qpn.ca/), an open-access cohort with extensive clinical characterization of participants with and without PD, were included.^40^ All iRBD patients received a video-polysomnography confirmed diagnosis based on the International Classification of Sleep Disorders, third edition (see Supplementary Material for criteria).^41^ The absence of concomitant DLB, PD and MSA was confirmed at the neurological evaluation closest in time to the neuroimaging acquisition. The exclusion criteria for all participants were: a) the presence of parkinsonism or dementia, b) a diagnosis of epilepsy or epileptiform abnormalities on EEG, c) antidepressant-induced RBD and d) the presence of sleep disorders mimicking RBD (sleepwalking, night terrors, uncontrolled sleep apnea). Most participants completed the Montreal Cognitive Assessment (MoCA)^42^ to evaluate global cognition and the Unified Parkinson’s Disease Rating Scale revised by MDS (MDS-UPDRS-III) scores,^43^ to evaluate severity of parkinsonian motor features. All participating centers received approval from their respective local research ethics committees and the multicentric project was approved by the Research Ethics Board of the CIUSSS du Nord-de-l’Île-de-Montréal and the McGill University Health Centre. All procedures adhered to research ethics guidelines and informed consent was obtained from each participant in accordance with the Declaration of Helsinki.

### Structural connectivity

Tractography and connectomic pipelines were applied to the DWI data to create structural connectivity matrices for each subject in the groups (Fig.6).

**Figure 6.**
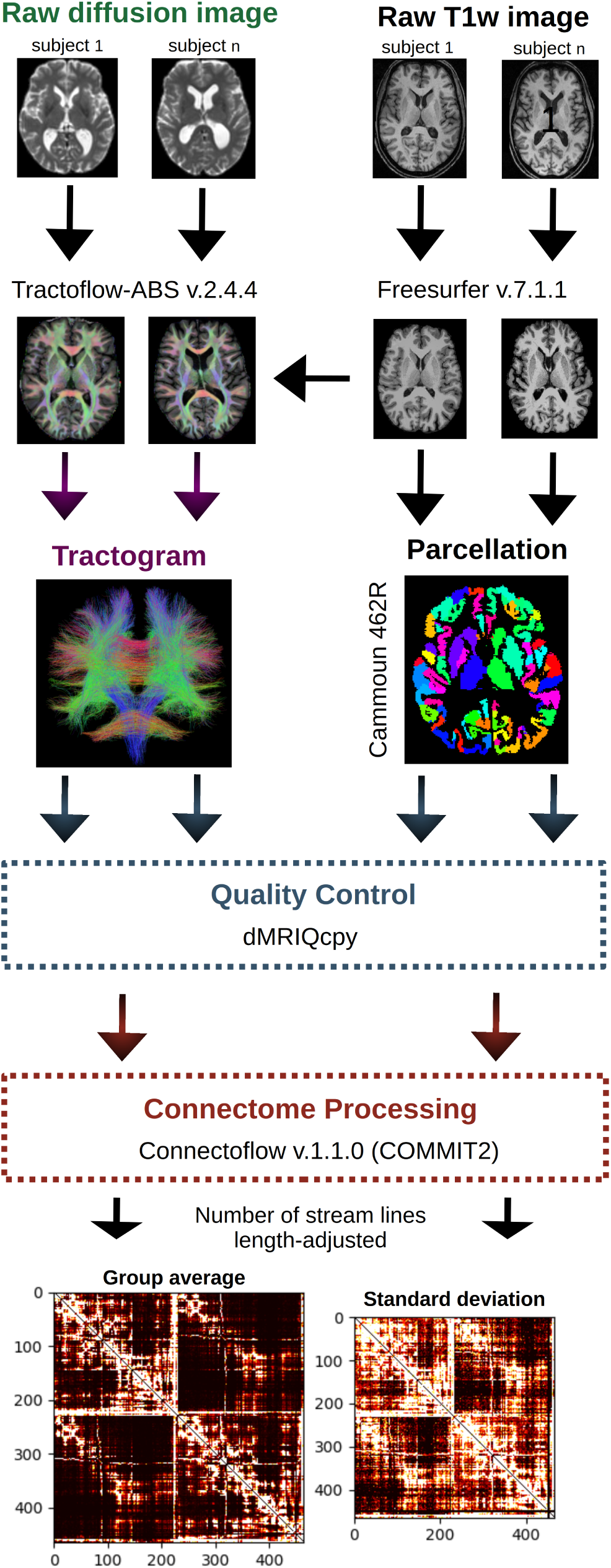
Main steps of the processing pipelines. The diffusion-weighted images (DWI) were first acquired from different cohorts with the T1-weighted images. The T1w processed with Freesurfer v.7.1.1 and DWI were used in Tractoflow-ABS v.2.4.4 to build a tractogram for each subject in the isolated REM sleep behavior disorder (iRBD) and healthy control groups. The Cammoun atlas with 448 cortical and 14 subcortical regions was registered in the T1w native space (ANTs registration) and quality control (dMRIQcpy) was performed for each main step of Tractoflow-ABS. Subsequently, the Connectoflow pipeline (v.1.1.0) was used with COMMIT2 to build a connectome for each subject (representing the number of streamlines, adjusted for the connection length, between each region of the atlas). The iRBD group average and standard deviation is shown for vizualisation.

### DWI processing

To generate structural connectivity matrices for each participant, we analyzed 592 DWI scans from all cohorts (289 iRBD and 303 controls). DWI scans were acquired between May 2012 and January 2023 using an echo-planar imaging sequence on a SIEMENS 3T scanner at each center. Acquisition parameters for DWI and T1w are described in the Supplementary Material. The TractoFlow Atlas-Based Segmentation pipeline (TractoFlow-ABS v.2.4.4: github.com/scilus/tractoflow),^44,45^ with the connectomics profile, was used to create tractograms from the raw DWI scans and T1w images resampled with FreeSurfer v.7.1.1 (processing is detailed in Supplementary Material).^46^ Using dMRIQCpy, we next produced quality control files for each main step of the tractography process to identify and remove DWI and T1w scans with artifacts.^47^ Thirteen subjects with iRBD and four controls (2.9% of scans) failed the Tractoflow-ABS processing. Additionally, 78 DWI scans from the iRBD group and 72 from the control group (26% of scans) were excluded following quality control due to motion artifacts, incomplete cortical or subcortical structures, inadequate correction of image distortions, inaccurate registration or incomplete tractogram data. This yielded a total of 425 scans for connectome reconstruction (Fig.7).

**Figure 7.**
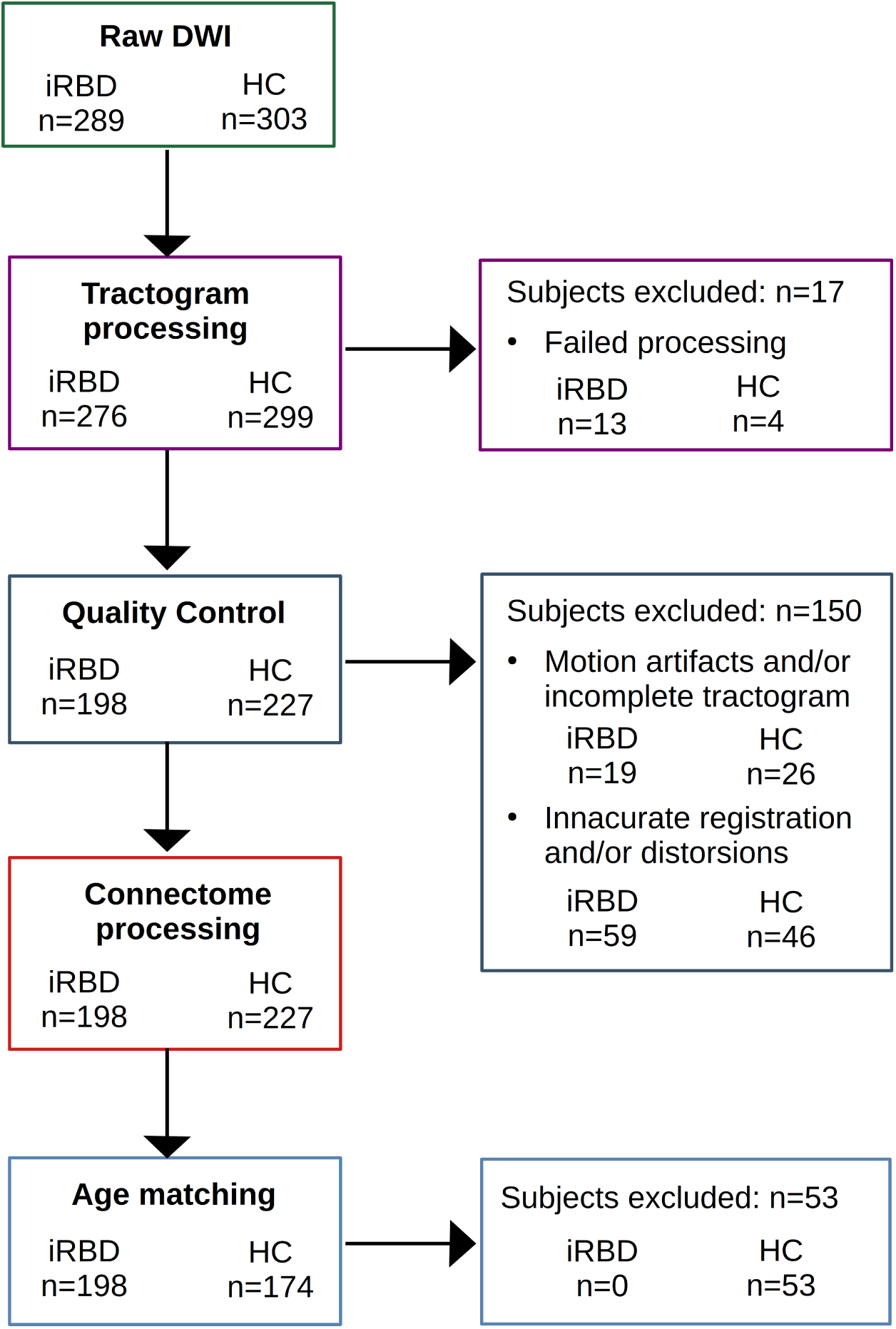
Flow chart showing the number of subjects in the isolated REM sleep behavior disorder (iRBD) and control (HC) groups at each main step of the method and exclusion criteria.

#### Structural connectome derivation and age matching

We used Connectoflow v.1.1.0 (github.com/scilus/connectoflow) to reconstruct structural connectomes for each participant.^48–50^ The connectome was built using the Cammoun atlas to obtain the number of streamlines (length-adjusted) between 448 cortical and 14 subcortical regions.^51^ First, the atlas was registered in the native space of each subject’s T1w MRI using ANTs (github.com/ANTsX/ANTs/).^52^ Tractograms generated by Tractoflow-ABS were processed by Connectoflow to generate whole-brain connectome. The COMMIT2 tool was applied to assign streamline weights and remove false positive connections.^21^ Group similarity matrices were built to exclude connections with extreme values in each subject’s connectivity matrix (with dissimilarity scores >12). Only intra-hemispheric connections were retained for analysis to remove potential artifacts since inter-hemispheric connections frequently involve complex fiber crossings that are inaccurately resolved using tractography.^53^ Connectivity values were transformed using the reciprocal of log10 to improve normality and interpretability.^54^ As a result, structural connectivity matrices were obtained for 198 iRBD patients and 227 controls (no subjects failed Connectoflow).

To minimize age-related confounding effects, younger controls (<55 years old) were excluded, aligning with the typical age of iRBD onset.^55^ This resulted in similar mean ages between the groups (iRBD mean (SD): 66.5Y (7.0); control: 66.2Y (6.9); *p-value* = 0.64). Reflecting the demographic trends in this population, the iRBD group had a higher men proportion (174 men/24 women) compared to the control group (99/75).^56^. The limited number of women, particularly in the iRBD group, prevented stratified analyses to assess sex effects, as such analyses would lack sufficient statistical power. To minimize potential effect, sex was controlled for in all analyses. In total, the structural connectivity matrices from 198 iRBD and 174 controls were available for analysis (Fig.7).

### Network-based statistics

The Network-Based Statistic toolbox v.1.2 (www.nitrc.org/projects/nbs/) was next used to compare the structural connectivity matrices between the iRBD and control groups, and to identify specific connections significantly affected in iRBD. Network-based statistics are a widely recognized method for group-level statistical analysis of structural and functional connectomes.^57^ This approach increases statistical power by considering interconnected sets of regions forming networks. To increase sensitivity to focal effects and avoid using arbitrary primary test statistic thresholds, we applied the false discovery rate (FDR) method (*p-value_FDR_* < 0.05, 10,000 permutations).^58^ Group was used as the independent variable, with age, sex, and center included as covariates in the design matrix.

### Graph theory metrics

We next determined whether network properties differed between iRBD and control connectomes. We used graph theory metrics, such as global and local efficiency, since these metrics have been particularly useful to identify early changes in brain networks due to pathology.^5^ We also investigated how local efficiency alterations relate to other topological features, namely clustering coefficient, connection strength, degree, and eigenvector centrality.^59^ Clustering coefficient measures how strongly regions in a network is connected to the nearby regions. Connection strength quantifies the density of connections between regions, degree refers to the total number of connections a region has, and eigenvector centrality measures a region’s influence based on both the quality (strength) and quantity (degree) of its connections. Supplementary Table 8 illustrates clustering coefficient and local efficiency patterns.

Specifically, global and local efficiency of the structural connectomes were calculated using the Brain Connectivity Toolbox (https://sites.google.com/site/bctnet) for each connectome (global efficiency) and each region of the Cammoun atlas (local efficiency).^59^ To account for inter-scanner variability, ComBat (github.com/Jfortin1/ComBatHarmonization), widely used in neuroimaging (including diffusion MRI),^17,60,61^ was applied on global and local efficiency measurements. Age and sex variance were preserved in ComBat. Regional W-scores were next calculated for each iRBD patient to regress out age and sex effects using the control group as reference.^62,63^ Linear statistical methods, specifically one-sample t-tests with FDR correction, were applied to the W-scores for global and local efficiency measures to identify significant differences between groups (*p-value_FDR_*< 0.05). A stepwise regression analysis was also conducted to identify which regions, in terms of local efficiency W-scores, were retained as significant predictors of global efficiency W-scores in iRBD patients.

Additional metrics potentially influencing local efficiency, such as clustering coefficient, connection strength, degree and eigenvector centrality, were computed using the Brain Connectivity toolbox.^59^ These measurements were scanner-corrected using ComBat^60^ and W-scored to regress out age and sex effects associated with normal aging.^62,63^ One sample t-tests (FDR-corrected) were applied on the W-scores for each metric to identify regions with significant difference between groups (*p-value_FDR_*< 0.05). Next, spatial Pearson correlations were calculated between local efficiency and each metric’s W-score. To account for spatial autocorrelation (i.e., the tendency of anatomically adjacent brain regions to have more similar measurements),^64^ correlations were compared to spatially constrained null models using the BrainSMASH toolbox^65^ (1000 spins, one-tailed, FDR-corrected). Additionally, to assess the differences between regions showing significantly higher and lower local efficiency in iRBD, we grouped these regions into two categories (positive and negative W-scores). We then compared their clustering coefficient, connection strength, degree, and eigenvector centrality using paired samples t-tests with FDR correction.

### Association with clinical features

The association between local efficiency and clinical features in iRBD was examined using the MoCA^42^ and MDS-UPDRS-III scores.^43^ Partial least squares (PLS) regression^66^ with 10 components was applied to identify latent variables maximizing the covariance between the local efficiency of the 462 brain regions (absolute W-scores representing deviations from the control group) used as dependent variables, and either the MoCA or MDS-UPDRS-III scores (Z-scores normalized to controls means) used as independent variables. The significance of each PLS latent variable was assessed by comparing it to a null distribution generated from 1000 random permutations. For each significant latent variable, the contribution of each region’s local efficiency to the latent variable was quantified, and bootstrapping (1000 iterations) was used to evaluate the significance of these contributions. Regions with a bootstrapping ratio exceeding 4.0 (indicating robust positive correlations between loading on the latent variable and clinical scores at *p-value* < 0.0001) or less than 4.0 (indicating robust negative correlations at *p-value* < 0.0001) were considered to significantly contribute to the latent variable. Only participants with iRBD and MoCA (N=186) or MDS-UPDRS-III scores (N=175) were included in the PLS regression.

### Relationship between local efficiency and phenoconversion status

Finally, we examined whether local efficiency (W-scores) was associated with phenoconversion in 177 iRBD patients under longitudinal follow-up. Among these patients, 43 (24.3%) had converted at the latest clinical visit: 28 (65.1%) converted to PD, 11 (25.6%) to DLB, and 4 (9.3%) to MSA. The remaining 134 individuals (75.7%) had not yet converted (non-converters). Patients who converted to MSA (n=4) were excluded due to their low number. The non-converters and, PD and DLB converters had similar age (F = 0.67, *p-value* = 0.51, Supplementary Fig.4), sex proportion (Fisher’s exact test *p-value* = 0.07), and time to phenoconversion (for converters, t = -1.23, *p-value* = 0.23, Supplementary Fig. 5). However, the non-converters had a longer follow-up time duration since MRI (6.9 years, SD = 3.0) compared with PD (t = 5.26, *p-value* < 0.0001) and DLB converters (t = 2.65, *p-value* = 0.02, Supplementary Fig. 5).

The predictive value of local efficiency (W-scores) for phenoconversion to PD or DLB was assessed across 462 regions. First, linear regression models (FDR-corrected) compared local efficiency between converters (to PD or DLB) and non-converters (remaining disease-free), adjusting for age, sex, and follow-up duration since MRI. Next, Cox proportional hazards regression analyses were conducted on regions with significant differences, controlling for age and sex, to examine associations between local efficiency and time to phenoconversion. Time zero was defined as MRI date. Survival time was the interval between MRI and the date of clinical visit confirming disease diagnosis (converters) or last follow-up (non-converters). Hazard ratios with 95% confidence intervals were reported. Kaplan–Meier survival curves were generated to compare the proportions of non-converters and converters between patients with positive versus negative local efficiency W-scores in significant regions.

## Data availability

The data used in this study were obtained from multiple collaborating centers, each of which retains ownership of their respective datasets. The principal investigator had authorized access to all data necessary for the analyses performed in this study. However, the accessibility and sharing of data are subject to the local policies and restriction criteria of each center involved. As such, data availability is restricted, and requests for access should be directed to the respective institutions, pending their specific data access and sharing guidelines. DWI data from the PPMI are publicly available at www.ppmi-info.org. The average structural connectivity matrices for the iRBD and control groups are available from the authors upon reasonable request. The software used can be accessed from the sources cited in the Methods section.

## Acknowledgements

We would like to thank Paul Cuciureanu for his help with the DWI data processing. J.F. Gagnon reports grants from the Fonds de recherche du Québec – Santé, the Canadian Institutes of Health Research, the W. Garfield Weston Foundation, the Michael J. Fox Foundation for Parkinson’s Research, and the National Institutes of Health, and holds a Canada Research Chair in Cognitive Decline in Pathological Aging. R. B. Postuma received grants from the CIHR, Michael J. Fox Foundation, NIH, Roche Diagnostics, and the Weston Foundation. He received consulting fees from Novartis, Eisai, Merck, Vaxxinity, BMS, Ventus, Korro, Vanqua, Roche, Regeneron, Helicon, Epic, and Clinilabs. He holds leadership roles with Parkinson Canada, the Michael J. Fox Foundation, MDS, Movement Disorders journal, and the RBD Study Group. P. Dusek, S. Marecek, and Z. Varga acknowledge support from the Czech Health Research Council (grant NU21-04-00535) and the National Institute for Neurological Research (Project No. LX22NPO5107), funded by the European Union – Next Generation EU, all paid to their institution. J. C. Klein acknowledges salary support from the National Institute for Health and Care Research (NIHR) Oxford Health Clinical Research Facility and the NIHR Oxford Biomedical Research Centre. S. Lehericy acknowledges support from the Programme d’investissements d’avenir (ANR-10-IAIHU-06), the Paris Institute of Neurosciences – IHU (IAIHU-06), Agence Nationale de la Recherche (ANR-11-INBS-0006), and Control-PD (JPND Cognitive Propagation in Prodromal Parkinson’s Disease), with payments made to institution. M. Hu acknowledges support from Parkinson’s UK, the Oxford Biomedical Research Centre, CPT, EPND, and the Michael J. Fox Foundation. S. Rahayel received grant support and travel reimbursement from the Michael J. Fox Foundation.

The Parkinson’s Progression Markers Initiative (PPMI) - a public-private partnership - is funded by the Michael J. Fox Foundation for Parkinson’s Research and funding partners, including 4D Pharma, AbbVie Inc., AcureX Therapeutics, Allergan, Amathus Therapeutics, Aligning Science Across Parkinson’s (ASAP), Avid Radiopharmaceuticals, Bial Biotech, Biogen, BioLegend, Bristol Myers Squibb, Calico Life Sciences LLC, Celgene Corporation, DaCapo Brainscience, Denali Therapeutics, The Edmond J. Safra Foundation, Eli Lilly and Company, GE Healthcare, GlaxoSmithKline, Golub Capital, Handl Therapeutics, Insitro, Janssen Pharmaceuticals, Lundbeck, Merck & Co., Inc., Meso Scale Diagnostics, LLC, Neurocrine Biosciences, Pfizer Inc., Piramal Imaging, Prevail Therapeutics, F. Hoffmann-La Roche Ltd and its affiliated company Genentech Inc., Sanofi Genzyme, Servier, Takeda Pharmaceutical Company, Teva Neuroscience, Inc., UCB, Vanqua Bio, Verily Life Sciences, Voyager Therapeutics, Inc., and Yumanity Therapeutics, Inc. For up-to-date information on the study, visit www.ppmi-info.org.

## Supplementary material

Supplementary material is available online.

## Appendix 1

List of the contributors involved in the ICEBERG Study Group:

Steering committee: Marie Vidailhet, MD, PhD, (Pitié-Salpêtrière Hospital, Paris, principal investigator of ICEBERG), Jean-Christophe Corvol, MD, PhD (Pitié-Salpêtrière Hospital, Paris, scientific lead), Isabelle Arnulf, MD, PhD (Pitié-Salpêtrière Hospital, Paris, member of the steering committee), Stéphane Lehericy, MD, PhD (Pitié-Salpêtrière Hospital, Paris, member of the steering committee);

Clinical data: Marie Vidailhet, MD, PhD, (Pitié-Salpêtrière Hospital, Paris, coordination), Graziella Mangone, MD, PhD (Pitié-Salpêtrière Hospital, Paris, co-coordination), Jean-Christophe Corvol, MD, PhD (Pitié-Salpêtrière Hospital, Paris), Isabelle Arnulf, MD, PhD (Pitié-Salpêtrière Hospital, Paris), Smaranda Leu MD (Pitié-Salpêtrière Hospital, Paris), Sara Sambin, MD (Pitié-Salpêtrière Hospital, Paris), Jonas Ihle, MD (Pitié-Salpêtrière Hospital, Paris), Caroline Weill, MD, (Pitié-Salpêtrière Hospital, Paris), Poornima MENON MD, (Pitié-Salpêtrière Hospital, Paris), David Grabli, MD, PhD (Pitié-Salpêtrière Hospital, Paris); Florence Cormier-Dequaire, MD (Pitié-Salpêtrière Hospital, Paris); Louise Laure Mariani, MD, PhD (Pitié-Salpêtrière Hospital, Paris), Emmanuel Roze, MD, PhD, (Pitié-Salpêtrière Hospital, Paris), Cécile Delorme, MD (Pitié-Salpêtrière Hospital, Paris), Elodie Hainque MD, PhD, (Pitié-Salpêtrière Hospital, Paris), Aurelie Méneret (MD, PhD, (Pitié-Salpêtrière Hospital, Paris), Bertrand Degos, MD, PhD (Avicenne Hospital, Bobigny);

Neuropsychological data: Richard Levy, MD (Pitié-Salpêtrière Hospital, Paris, coordination), Fanny Pineau, MS (Pitié-Salpêtrière Hospital, Paris, neuropsychologist), Julie Socha, MS (Pitié-Salpêtrière Hospital, Paris, neuropsychologist), Eve Benchetrit, MS (La Timone Hospital, Marseille, neuropsychologist), Virginie Czernecki, MS (Pitié-Salpêtrière Hospital, Paris, neuropsychologist), Marie-Alexandrine Glachant, MS (Pitié-Salpêtrière Hospital, Paris, neuropsychologist);

Eye movement: Sophie Rivaud-Pechoux, PhD (ICM, Paris, coordination); Elodie Hainque, MD, PhD (Pitié-Salpêtrière Hospital, Paris);

Sleep assessment: Isabelle Arnulf, MD, PhD (Pitié-Salpêtrière Hospital, Paris, coordination), Smaranda Leu Semenescu, MD (Pitié-Salpêtrière Hospital, Paris), Pauline Dodet, MD (Pitié-Salpêtrière Hospital, Paris);

Genetic data: Jean-Christophe Corvol, MD, PhD (Pitié-Salpêtrière Hospital, Paris, coordination), Graziella Mangone, MD, PhD (Pitié-Salpêtrière Hospital, Paris, co-coordination), Samir Bekadar, MS (Pitié-Salpêtrière Hospital, Paris, biostatistician), Alexis Brice, MD (ICM, Pitié-Salpêtrière Hospital, Paris), Suzanne Lesage, PhD (INSERM, ICM, Paris, genetic analyses);

Metabolomics: Fanny Mochel, MD, PhD (Pitié-Salpêtrière Hospital, Paris, coordination), Farid Ichou, PhD (ICAN, Pitié-Salpêtrière Hospital, Paris), Vincent Perlbarg, PhD, Pierre and Marie Curie University), Benoit Colsch, PhD (CEA, Saclay), Arthur Tenenhaus, PhD (Supelec, Gif-sur-Yvette, data integration);

Brain MRI data: Stéphane Lehericy, MD, PhD (Pitié-Salpêtrière Hospital, Paris, coordination), Rahul Gaurav, MS, (Pitié-Salpêtrière Hospital, Paris, data analysis), Nadya Pyatigorskaya, MD, PhD, (Pitié-Salpêtrière Hospital, Paris, data analysis); Lydia Yahia-Cherif, PhD (ICM, Paris, Biostatistics), Romain Valabregue, PhD (ICM, Paris, data analysis), Cécile Galléa, PhD (ICM, Paris);

DaTscan imaging data: Marie-Odile Habert, MCU-PH (Pitié-Salpêtrière Hospital, Paris, coordination);

Voice recording: Dijana Petrovska, PhD (Telecom Sud Paris, Evry, coordination), Laetitia Jeancolas, MS (Telecom Sud Paris, Evry);

Study management: Vanessa Brochard (Pitié-Salpêtrière Hospital, Paris, coordination), Alizé Chalançon (Pitié-Salpêtrière Hospital, Paris, Project manager), Carole Dongmo-Kenfack (Pitié-Salpêtrière Hospital, Paris, clinical research assistant); Christelle Laganot (Pitié-Salpêtrière Hospital, Paris, clinical research assistant), Valentine Maheo (Pitié-Salpêtrière Hospital, Paris, clinical research assistant), Manon Gomes (Pitié-Salpêtrière Hospital, Paris, clinical research assistant).

