## Supplementary Material for "Altered Network Efficiency in Isolated REM Sleep Behaviour Disorder: A Multicentric Study"

### *Supplementary Information*

#### International Classification of Sleep Disorders (third edition) criteria for the iRBD diagnosis

The diagnosis for isolated REM Sleep Behavior Disorder (iRBD) requires: a) repeated episodes of complex motor behaviors or vocalizations during sleep, b) confirmation by polysomnography (PSG) that these behaviors occur during REM sleep, or a clinical history supporting this, c) demonstration by PSG of loss of atonia during REM sleep, d) the absence of epileptiform activity during REM sleep, and e) that the manifestations are not better explained by another disorder.

### Acquisition parameters for the DWI scans

The parameters and number of scans before quality control for the iRBD group (n=276) were as follows: b-values of 700 s/mm² (N=86) and 1000 s/mm² (N=190); gradient directions = 21 (N=11), 30 (N=74), 31 (N=34), 32 (N=34), 62 (N=73), 64 (N=41), and 65 (N=9); voxel size = 1.7 mm isotropic (N=45) and 2.0 mm isotropic (N=232); TR ranging from 6.3 to 14 s; and TE ranging from 59 to 101 ms. The control group (n=299) had comparable scan parameters: b-value of 700 s/mm² (N=94) and 1000 s/mm² (N=205); gradient directions = 21 (N=12), 30 (N=55), 31 (N=47), 32 (N=45), 62 (N=59), 64 (N=37), and 65 (N=44); voxel size = 1.7 mm isotropic (N=57) and 2.0 mm isotropic (N=242); TR ranging from 6.5 to 14 s; and TE ranging from 59 to 101 ms.

### Acquisition parameters for the T1w MRI

The acquisition parameters for the T1-weighted MRI acquisition, the scans from Montreal were acquired with an MPRAGE sequence with the following parameters: TR = 2300 ms; TE = 2.91 ms; flip angle = 9°; and voxel size = 1 mm³ isotropic; or with TR = 2300 ms; TE = 2.98 ms; flip angle = 9°; and voxel size = 1 mm³ isotropic. The scans from Prague were acquired with an MPRAGE sequence with: TR = 2200 ms; TE = 2.4 ms; flip angle = 8°; and voxel size = 1 mm³ isotropic. The ones from Oxford used MPRAGE with TR = 2040 ms; TE = 4.7 ms; flip angle = 8°; and voxel size = 1 mm³ isotropic. The scans from Paris were acquired with an MPRAGE sequence with: TR = 2300 ms; TE = 4.18 ms; flip angle = 9°; and voxel size = 1 mm³ isotropic; or MP2RAGE with: TR = 5000 ms; TE = 2.98 ms; flip angles = 4° and 5°; GRAPPA = 3; and voxel size = 1 mm³ isotropic. The acquisition parameters for the Parkinson's Progression Markers Initiative study (PPMI) are described in the MRI Technical Operations Manual:
www.ppmi-info.org/sites/default/files/docs/archives
PPMI2.0_MRI_TOM_Final_FullyExecuted_v2.0_20200807.pdf

### DWI processing with Tractoflow-ABS

DWI processing involved denoising, followed by topup, eddy-current, and N4 bias corrections to adjust for distortions and inhomogeneities.[1,2] After these corrections, the tensor model was fit to the data to extract diffusion metrics.[3] The following parameters used for diffusion processing included: DTI shells of 0, 700, and 1000; fODF shells of 0, 700, and 1000; local probabilistic tracking algorithm; white matter/grey matter interface as the local seeding mask type; 20 seeds per voxel; and a spherical harmonic (SH) order of 6 for datasets with fewer than 32 gradient directions and 8 for datasets with 32 or more directions. A higher SH order allows for a more accurate representation of complex diffusion patterns in the presence of a higher number of gradient directions.[4] The T1w resampled were registered in DWI native space.[5]

Table.1

**Regions with connectivity changes in individuals with isolated REM sleep behavior disorder (iRBD) compared to healthy controls (HC)**

| iRBD | Hemisphere | Region (source) | Target Region | T-stat |
|---|---|---|---|---|
| Lower density | Right | inferior parietal_7 | fusiform_6 | 4.70 |
| | Right | lateral occipital_5 | lingual_7 | 4.33 |
| | Right | inferior parietal_5 | fusiform_6 | 3.89 |
| | Right | posterior cingulate_1 | insula_7 | 3.79 |
| | Right | middle temporal_7 | superior temporal_7 | 3.73 |
| | Right | parstriangularis_3 | lateral occipital_10 | 3.70 |
| | Right | supramarginal_7 | Inferior temporal_3 | 3.65 |
| | Right | superior frontal_16 | superior temporal_11 | 3.55 |
| | Right | precentral_4 | fusiform_5 | 3.33 |
| | Left | precentral_16 | paracentral_1 | 4.04 |
| | Left | parsorbitalis_1 | rostral middle frontal_5 | 3.88 |
| | Left | superior frontal_17 | rostral anterior cingulate_1 | 3.88 |
| | Left | lateral orbitofrontal_6 | caudal anterior cingulate_2 | 3.75 |
| | Left | postcentral_8 | postcentral_13 | 3.48 |
| Higher density | Right | superior parietal_12 | inferior parietal_9 | 4.58 |
| | Right | superior parietal_13 | inferior parietal_9 | 3.79 |
| | Right | superior parietal_11 | inferior parietal_9 | 3.52 |
| | Left | medial orbitofrontal_5 | lingual_2 | 3.29 |

Table.2

**Regions with significant local efficiency changes in the isolated REM sleep behavior disorder (iRBD) group**

| iRBD | Hemisphere | Region | W-score | *P-value*$_{FDR}$ |
|---|---|---|---|---|
| Lower Local Eficiency | Right | precentral_12 | -0.33 | 0.002 |
| | Right | superior frontal_7 | -0.32 | 0.00006 |
| | Right | superior frontal_2 | -0.26 | 0.008 |
| | Right | paracentral_3 | -0.25 | 0.008 |
| | Right | inferior parietal_6 | -0.22 | 0.008 |
| | Right | middle temporal_6 | -0.22 | 0.03 |
| | Right | thalamus proper | -0.22 | 0.04 |
| | Right | lingual_2 | -0.20 | 0.04 |
| | Left | postcentral_5 | -0.52 | 0.0000000002 |
| | Left | putamen | -0.41 | 0.000001 |
| | Left | inferior parietal_4 | -0.41 | 0.00002 |
| | Left | thalamus proper | -0.37 | 0.00006 |
| | Left | supramarginal_4 | -0.31 | 0.0007 |
| | Left | middle temporal_1 | -0.27 | 0.004 |
| | Left | parsopercularis_4 | -0.26 | 0.01 |
| | Left | cuneus_1 | -0.26 | 0.01 |
| | Left | fusiform_6 | -0.26 | 0.008 |
| | Left | superior frontal_7 | -0.24 | 0.01 |
| | Left | middle temporal_3 | -0.22 | 0.01 |
| | Left | inferior parietal_9 | -0.22 | 0.01 |
| | Left | insula_3 | -0.21 | 0.03 |
| Higher Local Efficiency | Right | inferior temporal_4 | 0.35 | 0.001 |
| | Right | lateral occipital_4 | 0.29 | 0.002 |
| | Right | lateral orbitofrontal_2 | 0.26 | 0.009 |
| | Right | caudal middle frontal_3 | 0.25 | 0.01 |
| | Right | precentral_14 | 0.25 | 0.007 |
| | Right | precuneus_5 | 0.24 | 0.02 |
| | Left | superior parietal_12 | 0.29 | 0.006 |
| | Left | superior temporal_4 | 0.27 | 0.01 |
| | Left | superior frontal_15 | 0.26 | 0.008 |
| | Left | bankssts_1 | 0.23 | 0.04 |
| | Left | lateral orbitofrontal_6 | 0.22 | 0.04 |

Table.3

**Regions retained as predictors of global efficiency changes in isolated REM sleep behavior disorder**

| Hemisphere | Region | Estimate | T-stat | *P-value* |
|---|---|---|---|---|
| Right | thalamus proper | 0.30 | 4.17 | 0.00005 |
| Left | putamen | 0.28 | 4.09 | 0.00006 |
| Left | lateral orbitofrontal_6 | 0.27 | 4.30 | 0.00003 |
| Left | thalamus proper | 0.18 | 2.29 | 0.02 |
| Right | lateral orbitofrontal_2 | -0.15 | -2.60 | 0.01 |
| Left | insula_3 | -0.15 | -2.26 | 0.02 |
| Left | inferior parietal_9 | -0.14 | -1.99 | 0.048 |
| Left | superior parietal_12 | -0.12 | -2.16 | 0.03 |

Table.4

**Regions with significant clustering coefficients changes in the isolated REM sleep behavior disorder (iRBD) group**

| iRBD | Hemisphere | Region | W-score | *P-value*$_{FDR}$ |
|---|---|---|---|---|
| Lower clustering coefficient | Right | precentral_12 | -0.36 | 0.0002 |
| | Right | superior frontal_7 | -0.33 | 0.00002 |
| | Right | inferior parietal_9 | -0.29 | 0.004 |
| | Right | middle temporal_6 | -0.27 | 0.003 |
| | Right | thalamus proper | -0.26 | 0.004 |
| | Right | superior frontal_2 | -0.26 | 0.004 |
| | Right | paracentral_3 | -0.26 | 0.003 |
| | Right | supramarginal_6 | -0.25 | 0.004 |
| | Right | posterior cingulate_4 | -0.25 | 0.005 |
| | Right | inferior parietal_6 | -0.24 | 0.003 |
| | Right | isthmus cingulate_1 | -0.23 | 0.02 |
| | Right | caudate | -0.23 | 0.02 |
| | Right | pallidum | -0.22 | 0.02 |
| | Right | superior frontal_13 | -0.21 | 0.02 |
| | Right | precentral_11 | -0.21 | 0.03 |
| | Right | middle temporal_1 | -0.20 | 0.03 |
| | Right | accumbens area | -0.20 | 0.04 |
| | Right | putamen | -0.20 | 0.04 |
| | Right | parahippocampal_2 | -0.19 | 0.03 |
| | Right | lingual_5 | -0.18 | 0.04 |
| | Left | postcentral_5 | -0.57 | $3x10^{-12}$ |
| | Left | inferior parietal_4 | -0.42 | 0.00001 |
| | Left | thalamus proper | -0.42 | 0.000001 |
| | Left | putamen | -0.39 | 0.0000009 |
| | Left | supramarginal_4 | -0.34 | 0.00006 |
| | Left | lateral occipital_4 | -0.33 | 0.001 |
| | Left | parsopercularis_4 | -0.31 | 0.001 |
| | Left | fusiform_6 | -0.29 | 0.001 |
| | Left | superior frontal_7 | -0.28 | 0.001 |
| | Left | middle temporal_1 | -0.28 | 0.001 |
| | Left | precuneus_11 | -0.27 | 0.01 |
| | Left | caudate | -0.26 | 0.003 |
| | Left | cuneus_1 | -0.26 | 0.008 |
| | Left | hippocampus | -0.26 | 0.02 |

| | | | | |
|---|---|---|---|---|
| | Left | inferior parietal_10 | -0.24 | 0.02 |
| | Left | middle temporal_3 | -0.24 | 0.004 |
| | Left | inferior parietal_9 | -0.23 | 0.004 |
| | Left | transverse temporal_1 | -0.23 | 0.007 |
| | Left | insula_3 | -0.22 | 0.009 |
| | Left | parahippocampal_3 | -0.22 | 0.04 |
| | Left | superior temporal_2 | -0.21 | 0.03 |
| | Left | supramarginal_7 | -0.21 | 0.03 |
| | Left | isthmus cingulate_1 | -0.20 | 0.03 |
| | Left | superior temporal_6 | -0.20 | 0.03 |
| | Left | supramarginal_10 | -0.19 | 0.04 |
| | Left | inferior temporal_5 | -0.19 | 0.02 |
| | Left | supramarginal_1 | -0.19 | 0.02 |
| | Left | precuneus_3 | -0.19 | 0.02 |
| Higher clustering coefficient | Right | inferior temporal_4 | 0.37 | 0.0006 |
| | Right | lateral occipital_4 | 0.29 | 0.001 |
| | Right | lateral orbitofrontal_2 | 0.23 | 0.02 |
| | Right | precentral_14 | 0.21 | 0.03 |
| | Right | fusiform_5 | 0.21 | 0.03 |
| | Left | superior parietal_12 | 0.32 | 0.001 |
| | Left | lateral orbitofrontal_6 | 0.25 | 0.008 |
| | Left | superior frontal_15 | 0.24 | 0.02 |
| | Left | superior temporal_4 | 0.24 | 0.02 |

Table.5

**Regions with significant connection strength changes in the isolated REM sleep behavior disorder (iRBD) group**

| iRBD | Hemisphere | Region | W-score | *P-value*$_{FDR}$ |
|---|---|---|---|---|
| Lower connection strength | Right | parsopercularis_2 | -0.40 | 3x10-6 |
| | Right | superior parietal_6 | -0.38 | 0.0002 |
| | Right | inferior temporal_4 | -0.35 | 0.0001 |
| | Right | lateral orbitofrontal_2 | -0.35 | 6x10-5 |
| | Right | lateral occipital_4 | -0.35 | 1x10-4 |
| | Right | precentral_14 | -0.34 | 5x10-5 |
| | Right | postcentral_5 | -0.32 | 0.0004 |
| | Right | superior frontal_6 | -0.31 | 0.002 |
| | Right | rostral middle frontal_12 | -0.31 | 8x10-5 |
| | Right | superior frontal_12 | -0.29 | 0.0001 |
| | Right | caudal middle frontal_3 | -0.28 | 0.005 |
| | Right | lateral occipital_7 | -0.27 | 0.006 |
| | Right | putamen | -0.26 | 0.01 |
| | Right | rostral middle frontal_5 | -0.26 | 0.005 |
| | Right | superior parietal_5 | -0.26 | 0.006 |
| | Right | superior parietal_4 | -0.26 | 0.009 |
| | Right | parstriangularis_1 | -0.25 | 0.006 |
| | Right | superior parietal_7 | -0.24 | 0.01 |
| | Right | posterior cingulate_1 | -0.23 | 0.006 |
| | Right | inferior parietal_5 | -0.23 | 0.03 |
| | Right | insula_6 | -0.23 | 0.01 |
| | Right | superior temporal_11 | -0.23 | 0.02 |
| | Right | caudal anterior cingulate_2 | -0.23 | 0.01 |
| | Right | inferior parietal_1 | -0.23 | 0.03 |
| | Right | superior frontal_1 | -0.23 | 0.009 |
| | Right | superior frontal_15 | -0.22 | 0.009 |
| | Right | precentral_5 | -0.22 | 0.01 |
| | Right | rostral middle frontal_8 | -0.22 | 0.02 |
| | Right | fusiform_5 | -0.22 | 0.02 |
| | Right | insula_2 | -0.22 | 0.02 |
| | Right | caudal middle frontal_4 | -0.22 | 0.01 |
| | Right | superior parietal_11 | -0.21 | 0.009 |
| | Right | superior frontal_11 | -0.21 | 0.01 |
| | Right | postcentral_12 | -0.21 | 0.02 |
| | Right | precentral_13 | -0.21 | 0.03 |

| | | | |
|---|---|---|---|
| Right | rostral middle frontal_2 | -0.2 | 0.01 |
| Right | supramarginal_1 | -0.2 | 0.02 |
| Right | fusiform_6 | -0.2 | 0.03 |
| Right | caudal anterior cingulate_1 | -0.2 | 0.02 |
| Right | bankssts_1 | -0.2 | 0.049 |
| Right | inferior temporal_3 | -0.19 | 0.04 |
| Right | superior temporal_2 | -0.19 | 0.04 |
| Right | superior temporal_10 | -0.19 | 0.03 |
| Right | postcentral_7 | -0.18 | 0.03 |
| Right | precentral_7 | -0.18 | 0.0497 |
| Right | insula_7 | -0.18 | 0.045 |
| Right | superior frontal_8 | -0.18 | 0.04 |
| Right | precentral_4 | -0.18 | 0.045 |
| Right | insula_4 | -0.18 | 0.04 |
| Right | amygdala | -0.16 | 0.04 |
| Left | parsopercularis_3 | -0.48 | $2x10^{-7}$ |
| Left | superior temporal_4 | -0.45 | $3x10^{-6}$ |
| Left | amygdala | -0.35 | $6x10^{-5}$ |
| Left | precentral_14 | -0.34 | $6x10^{-5}$ |
| Left | superior temporal_8 | -0.33 | $5x10^{-5}$ |
| Left | superior parietal_12 | -0.32 | 0.0001 |
| Left | lateral orbitofrontal_6 | -0.32 | $1x10^{-5}$ |
| Left | parstriangularis_1 | -0.31 | 0.0007 |
| Left | superior frontal_15 | -0.29 | 0.001 |
| Left | lateral orbitofrontal_4 | -0.29 | 0.001 |
| Left | superior frontal_16 | -0.28 | 0.0004 |
| Left | caudal middle frontal_3 | -0.28 | 0.0004 |
| Left | inferior parietal_8 | -0.28 | 0.01 |
| Left | rostral middle frontal_6 | -0.26 | 0.003 |
| Left | lingual_2 | -0.26 | 0.005 |
| Left | precentral_8 | -0.26 | 0.003 |
| Left | postcentral_12 | -0.25 | 0.003 |
| Left | parsopercularis_1 | -0.25 | 0.009 |
| Left | caudal middlef rontal_4 | -0.25 | 0.006 |
| Left | rostral anterior cingulate_1 | -0.25 | 0.006 |
| Left | precentral_15 | -0.24 | 0.02 |
| Left | isthmuscingulate_2 | -0.24 | 0.02 |
| Left | parsorbitalis_1 | -0.23 | 0.01 |
| Left | caudal middle frontal_6 | -0.23 | 0.0006 |
| Left | superior temporal_3 | -0.23 | 0.006 |

| | | | | |
|---|---|---|---|---|
| | Left | superior parietal_7 | -0.23 | 0.02 |
| | Left | putamen | -0.23 | 0.02 |
| | Left | superior frontal_17 | -0.23 | 0.01 |
| | Left | precentral_13 | -0.23 | 0.01 |
| | Left | posterior cingulate_3 | -0.23 | 0.02 |
| | Left | precentral_11 | -0.22 | 0.01 |
| | Left | inferior parietal_1 | -0.22 | 0.03 |
| | Left | superior parietal_6 | -0.22 | 0.01 |
| | Left | precentral_9 | -0.21 | 0.01 |
| | Left | inferior parietal_3 | -0.21 | 0.02 |
| | Left | rostral middle frontal_8 | -0.21 | 0.01 |
| | Left | lateral occipital_10 | -0.21 | 0.04 |
| | Left | superior frontal_6 | -0.21 | 0.04 |
| | Left | superior parietal_3 | -0.21 | 0.03 |
| | Left | superior temporal_7 | -0.21 | 0.03 |
| | Left | postcentral_2 | -0.2 | 0.04 |
| | Left | inferior parietal_7 | -0.2 | 0.04 |
| | Left | postcentral_7 | -0.2 | 0.02 |
| | Left | parstriangularis_2 | -0.19 | 0.03 |
| | Left | accumbens area | -0.19 | 0.04 |
| | Left | superior frontal_18 | -0.19 | 0.045 |
| | Left | precentral_12 | -0.19 | 0.02 |
| | Left | lingual_1 | -0.18 | 0.01 |
| | Left | bankssts_1 | -0.18 | 0.045 |
| | Left | precentral_2 | -0.17 | 0.0497 |
| | Left | precuneus_2 | -0.17 | 0.04 |
| | Left | caudal anterior cingulate_2 | -0.17 | 0.049 |
| Higher connection strength | Right | precentral_8 | 0.19 | 0.03 |
| | Right | superior frontal_7 | 0.20 | 0.02 |
| | Right | medialor bitofrontal_3 | 0.20 | 0.02 |
| | Right | inferior parietal_9 | 0.24 | 0.02 |
| | Right | lingual_5 | 0.24 | 0.02 |
| | Left | postcentral_5 | 0.30 | 0.0004 |
| | Left | fusiform_6 | 0.22 | 0.03 |

Table.6

**Regions with significant degree changes in the isolated REM sleep behavior disorder (iRBD) group**

| iRBD | Hemisphere | Region | W-score | *P-value*$_{FDR}$ |
|---|---|---|---|---|
| Lower Degree | Right | parsopercularis_2 | -0.40 | $2x10^{-6}$ |
| | Right | superior parietal_6 | -0.38 | 0.0001 |
| | Right | lateral occipital_4 | -0.36 | $7x10^{-5}$ |
| | Right | lateral orbitofrontal_2 | -0.36 | $7x10^{-5}$ |
| | Right | inferior temporal_4 | -0.35 | 0.0001 |
| | Right | precentral_14 | -0.34 | $7x10^{-5}$ |
| | Right | postcentral_5 | -0.32 | 0.001 |
| | Right | superior frontal_6 | -0.31 | 0.001 |
| | Right | rostral middle frontal_12 | -0.30 | 0.0001 |
| | Right | caudal middle frontal_3 | -0.29 | 0.003 |
| | Right | putamen | -0.28 | 0.008 |
| | Right | superior frontal_12 | -0.28 | 0.0004 |
| | Right | superior parietal_4 | -0.27 | 0.006 |
| | Right | superior parietal_5 | -0.27 | 0.004 |
| | Right | rostral middle frontal_5 | -0.26 | 0.004 |
| | Right | lateral occipital_7 | -0.26 | 0.008 |
| | Right | insula_6 | -0.25 | 0.006 |
| | Right | inferior parietal_1 | -0.25 | 0.02 |
| | Right | parstriangularis_1 | -0.25 | 0.008 |
| | Right | superior temporal_11 | -0.24 | 0.02 |
| | Right | posterior cingulate_1 | -0.24 | 0.007 |
| | Right | caudal middle frontal_4 | -0.23 | 0.009 |
| | Right | superiorparietal_7 | -0.23 | 0.01 |
| | Right | inferior parietal_5 | -0.23 | 0.03 |
| | Right | postcentral_12 | -0.23 | 0.01 |
| | Right | superior frontal_1 | -0.22 | 0.01 |
| | Right | insula_2 | -0.22 | 0.02 |
| | Right | fusiform_5 | -0.22 | 0.02 |
| | Right | rostral middle frontal_8 | -0.22 | 0.02 |
| | Right | precentral_5 | -0.22 | 0.01 |
| | Right | superior parietal_11 | -0.22 | 0.008 |
| | Right | caudal anterior cingulate_2 | -0.22 | 0.02 |
| | Right | superior frontal_15 | -0.21 | 0.01 |
| | Right | precentral_13 | -0.21 | 0.02 |
| | Right | caudal anterior cingulate_1 | -0.21 | 0.02 |
| | Right | rostral middle frontal_2 | -0.21 | 0.01 |

| | | | |
|---|---|---|---|
| Right | superior frontal_11 | -0.21 | 0.01 |
| Right | insula_7 | -0.20 | 0.03 |
| Right | fusiform_6 | -0.20 | 0.02 |
| Right | supramarginal_1 | -0.20 | 0.02 |
| Right | superior temporal_10 | -0.20 | 0.02 |
| Right | precentral_7 | -0.19 | 0.04 |
| Right | inferior temporal_3 | -0.19 | 0.04 |
| Right | superior temporal_2 | -0.19 | 0.04 |
| Right | superior frontal_10 | -0.18 | 0.04 |
| Right | insula_4 | -0.18 | 0.03 |
| Right | parsorbitalis_1 | -0.18 | 0.04 |
| Right | postcentral_7 | -0.18 | 0.03 |
| Right | parstriangularis_4 | -0.17 | 0.05 |
| Right | superior frontal_8 | -0.17 | 0.04 |
| Right | lateral occipital_3 | -0.17 | 0.04 |
| Right | accumbens area | -0.17 | 0.03 |
| Left | parsopercularis_3 | -0.48 | $3x10^{-7}$ |
| Left | superior temporal_4 | -0.45 | $2x10^{-6}$ |
| Left | precentral_14 | -0.35 | $3x10^{-5}$ |
| Left | amygdala | -0.34 | 0.0001 |
| Left | lateral orbitofrontal_6 | -0.32 | $1x10^{-5}$ |
| Left | superior parietal_12 | -0.32 | 0.0001 |
| Left | superior temporal_8 | -0.31 | $9x10^{-5}$ |
| Left | superior frontal_15 | -0.31 | 0.0005 |
| Left | parstriangularis_1 | -0.29 | 0.001 |
| Left | lateral orbitofrontal_4 | -0.29 | 0.001 |
| Left | caudal middle frontal_3 | -0.29 | 0.0003 |
| Left | superior frontal_16 | -0.28 | 0.001 |
| Left | inferior parietal_8 | -0.27 | 0.01 |
| Left | precentral_8 | -0.25 | 0.003 |
| Left | Isthmus cingulate_2 | -0.25 | 0.02 |
| Left | postcentral_12 | -0.25 | 0.004 |
| Left | posterior cingulate_3 | -0.24 | 0.01 |
| Left | lingual_2 | -0.24 | 0.007 |
| Left | parsopercularis_1 | -0.24 | 0.01 |
| Left | caudal middle frontal_4 | -0.24 | 0.008 |
| Left | rostral middle frontal_6 | -0.24 | 0.008 |
| Left | superior frontal_17 | -0.24 | 0.007 |
| Left | caudal middle frontal_6 | -0.23 | 0.001 |
| Left | rostral anterior cingulate_1 | -0.23 | 0.009 |

| | | | | |
|---|---|---|---|---|
| | Left | superior parietal_7 | -0.23 | 0.01 |
| | Left | precentral_15 | -0.23 | 0.02 |
| | Left | precentral_13 | -0.23 | 0.009 |
| | Left | superior parietal_6 | -0.22 | 0.007 |
| | Left | rostral middle frontal_8 | -0.22 | 0.01 |
| | Left | precentral_11 | -0.22 | 0.01 |
| | Left | inferior parietal_1 | -0.22 | 0.03 |
| | Left | superior temporal_3 | -0.22 | 0.009 |
| | Left | parsorbitalis_1 | -0.21 | 0.02 |
| | Left | superior frontal_6 | -0.21 | 0.04 |
| | Left | precentral_9 | -0.21 | 0.01 |
| | Left | inferior parietal_3 | -0.21 | 0.03 |
| | Left | lateral occipital_10 | -0.21 | 0.04 |
| | Left | postcentral_8 | -0.21 | 0.04999 |
| | Left | superior parietal_3 | -0.21 | 0.03 |
| | Left | putamen | -0.20 | 0.04 |
| | Left | superior temporal_7 | -0.20 | 0.03 |
| | Left | postcentral_2 | -0.20 | 0.04 |
| | Left | bankssts_1 | -0.20 | 0.03 |
| | Left | superior frontal_18 | -0.20 | 0.04 |
| | Left | precentral_12 | -0.20 | 0.01 |
| | Left | inferior parietal_7 | -0.20 | 0.04 |
| | Left | postcentral_7 | -0.19 | 0.02 |
| | Left | accumben sarea | -0.19 | 0.04 |
| | Left | parstriangularis_2 | -0.18 | 0.045 |
| | Left | lingual_1 | -0.18 | 0.01 |
| | Left | precuneus_2 | -0.18 | 0.03 |
| | Left | caudal anterior cingulate_2 | -0.18 | 0.04 |
| | Left | precentral_4 | -0.16 | 0.04 |
| Higher Degree | Right | lingual_5 | 0.26 | 0.01 |
| | Right | inferior parietal_9 | 0.22 | 0.03 |
| | Right | superior frontal_7 | 0.21 | 0.01 |
| | Right | precentral_8 | 0.18 | 0.04 |
| | Right | medial orbitofrontal_3 | 0.18 | 0.04999 |
| | Left | postcentral_5 | 0.30 | 0.0004 |
| | Left | fusiform_6 | 0.22 | 0.03 |

Table.7

**Number of subjects in each cohort at each step of the method for the isolated REM sleep behavior disorder (iRBD) and healthy control (HC) groups**

| Data | Group | Study/ Center | City (Country) | Raw data | Tractogram processing | QC | Connectome processing | Group balancing |
|---|---|---|---|---|---|---|---|---|
| DWI-MRI | iRBD | ALICE | Paris (France) | 14 | 11 | 4 | 4 | 4 |
| | | CRIUGM | Montreal (Canada) | 41 | 41 | 29 | 29 | 29 |
| | | ICEBERG | Paris (France) | 35 | 34 | 33 | 33 | 33 |
| | | Discovery | Oxford (UK) | 73 | 73 | 46 | 46 | 46 |
| | | PPMI | Multiple* | 9 | 9 | 6 | 6 | 6 |
| | | Roche | Montreal (Canada) | 34 | 34 | 34 | 34 | 34 |
| | | Prague | Prague (Czechia) | 83 | 74 | 46 | 46 | 46 |
| | | **Total** | - | **289** | **276** | **198** | **198** | **198** |
| | HC | ALICE | Paris (France) | 13 | 12 | 4 | 4 | 4 |
| | | CRIUGM | Montreal (Canada) | 37 | 37 | 29 | 29 | 24 |
| | | ICEBERG | Paris (France) | 45 | 45 | 40 | 40 | 31 |
| | | Discovery | Oxford (UK) | 59 | 59 | 30 | 30 | 26 |
| | | PPMI | Multiple* | 45 | 44 | 41 | 41 | 28 |
| | | QPN | Montreal (Canada) | 47 | 47 | 44 | 44 | 36 |
| | | Prague | Prague (Czechia) | 57 | 55 | 39 | 39 | 25 |
| | | **Total** | - | **303** | **299** | **227** | **227** | **174** |

*Participants recruited within the PPMI were recruited from the USA, Canada, Greece, Spain, Luxembourg, Nigeria, UK, Germany, Netherlands, and Israel

DWI-MRI = Diffusion-weighted magnetic resonance imaging; QC: Quality Control; PPMI = Parkinson’s Progression Markers Initiative; QPN = Quebec Parkinson Network

Table. 8

**Example of network configurations based on local efficiency and clustering coefficient**

| | Network example | |
|---|---|---|
| **High** local efficiency and clustering coefficient | A network where a region's neighbors are well-connected to each other (high clustering) and information can travel through multiple path between them (high local efficiency). | 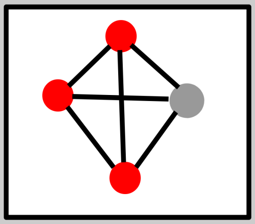 |
| **Low** local efficiency and clustering coefficient | A network where a region's neighbors are not well-connected to each other (low clustering), and information transfer is slow because it has to pass through a long path to reach a specific region in the network (low local efficiency). | 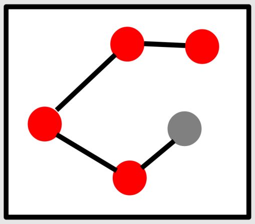 |

Figure 1. **Distribution of the global efficiency scores (W-score) in individuals with isolated REM sleep behavior disorder (iRBD)**

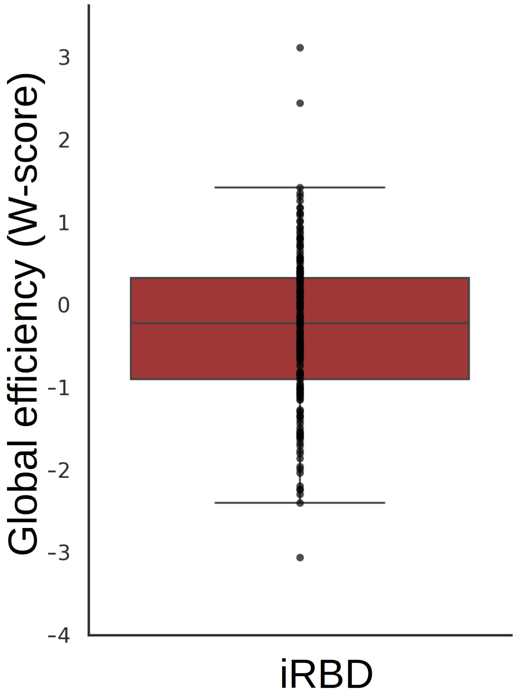

Figure 2. **Variance explained by each latent variable (A) and participants with isolated REM sleep behavior disorder (iRBD) loading for the third latent variable (B)**

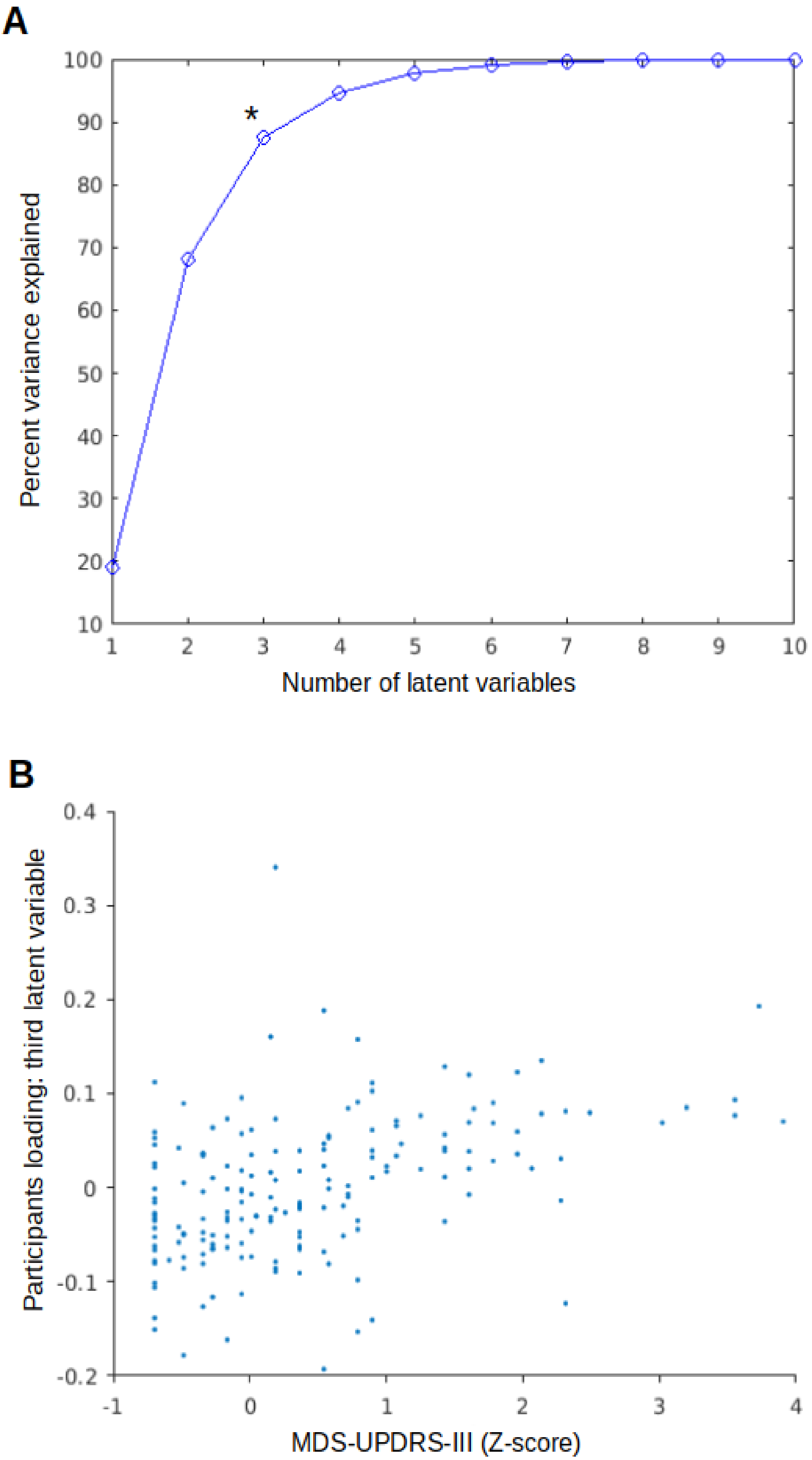

Figure 3. **Relationships with parkinsonian motor features in isolated REM sleep behavior disorder (iRBD)**

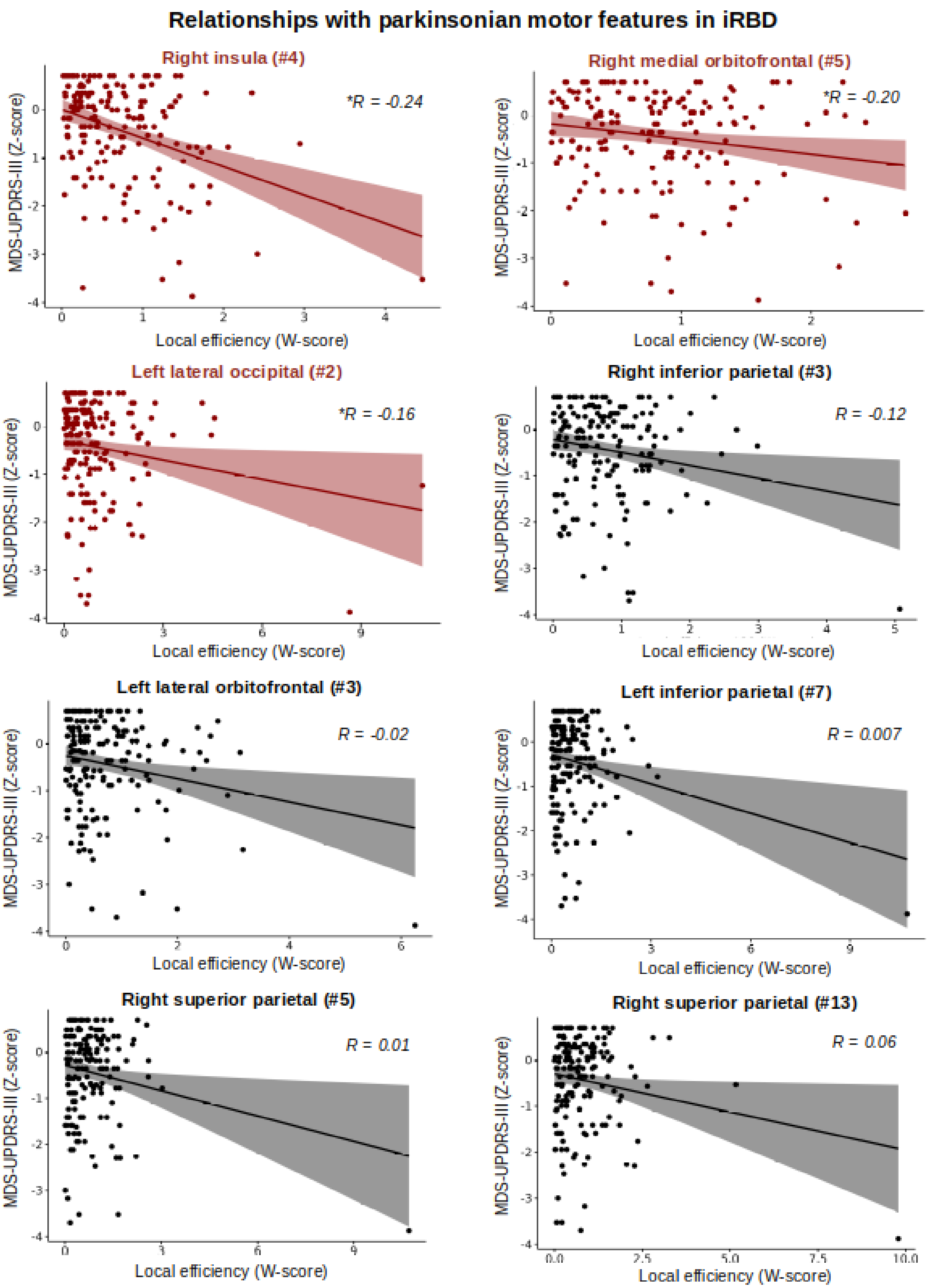

Figure 4. **Similar age distributions for the individuals with iRBD who had not yet converted (non-converters. N = 134) and the iRBD patients who converted to PD (N=28) and DLB N=11)**

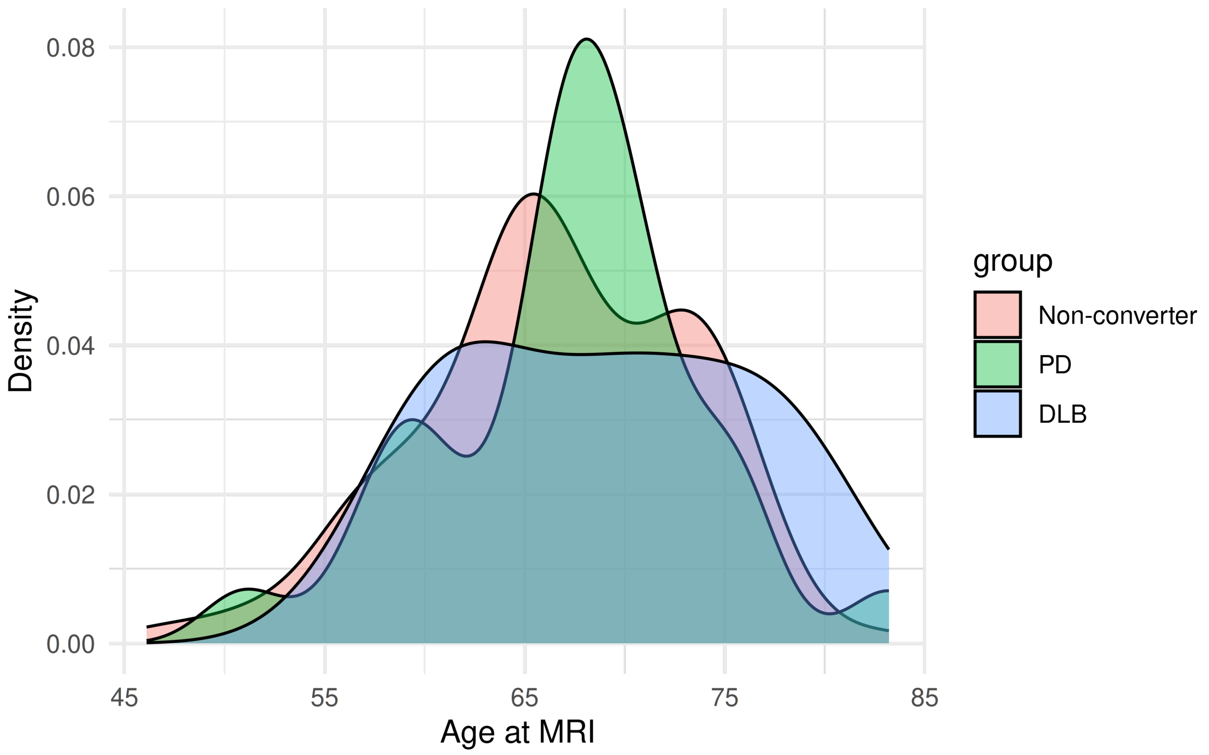

Figure 5. **Similar follow-up time for the individuals with iRBD who had not yet converted (non-converters. N = 134) and the iRBD patients who converted to PD (N=28) and DLB N=11)**

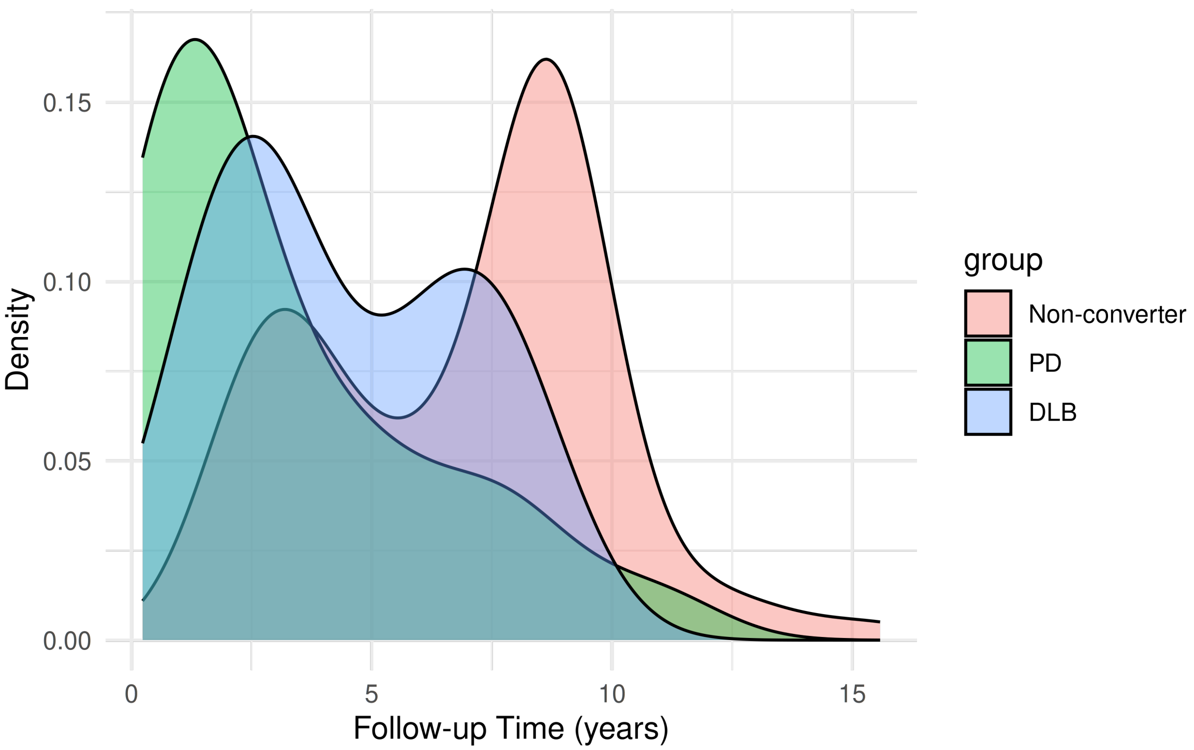
